# The genetic and phenotypic correlates of mtDNA copy number in a multi-ancestry cohort

**DOI:** 10.1101/2022.10.21.22281387

**Authors:** Arslan A. Zaidi, Anurag Verma, Colleen Morse, Penn Medicine BioBank, Marylyn D. Ritchie, Iain Mathieson

## Abstract

Mitochondrial DNA copy number (mtCN) is often treated as a proxy for mitochondrial (dys)function and disease risk. Pathological changes in mtCN are common symptoms of rare mitochondrial disorders but reported associations between mtCN and common diseases vary considerably across studies. We sought to understand the biology of mtCN by carrying out genome and phenome-wide association studies of mtCN in 30,666 individuals from the Penn Medicine BioBank—a large, diverse cohort of largely African and European ancestry. We estimated mtCN in peripheral blood using exome sequence data, taking into account the effects of blood cell composition, particularly neutrophil and platelet counts. We replicated known genetic associations of mtCN in the PMBB and found that their effect sizes are highly correlated between individuals of European and African ancestry. However, the heritability of mtCN was much higher among individuals of largely African ancestry (*h*^2^ = 0.3) compared to European ancestry individuals (*h*^2^ = 0.1). Admixture mapping suggests that there are undiscovered variants underlying mtCN that are differentiated in frequency between individuals with African and European ancestry. We further show that mtCN is associated with many health-related phenotypes. We discovered robust associations between mtDNA copy number and diseases of metabolically active tissues, such as cardiovascular disease and liver damage that were consistent across African and European ancestry individuals. Other associations, such as epilepsy, prostate cancer, and disorders of iron metabolism were only discovered in either individuals with European or African ancestry, but not both. Even though we replicate known genetic and phenotypic associations of mtCN, we demonstrate that they are sensitive to blood cell composition and environmental modifiers, explaining why such associations are inconsistent across studies. Peripheral blood mtCN might therefore be used as a biomarker of mitochondrial dysfunction and disease risk, but such associations must be interpreted with care.

## Introduction

Mitochondria are vital to cellular function, playing important roles in energy production, calcium signaling, cellular homeostasis, apoptosis and synthesis of biomolecules. Mitochondrial function is mediated by more than 1,000 proteins—of which only 13 are encoded by the mitochondrial DNA (mtDNA), with the rest encoded by the nuclear genome [1]. Loss of function mutations in these genes can lead to mitochondrial dysfunction, which typically affects multiple systems and tends to be clinically heterogeneous [2]. Considerable effort has been made to understand the genetics of mitochondrial dysfunction through family-based studies of rare mitochondrial diseases [3]. But, the extent to which mitochondrial dysfunction contributes to, or is affected by, common diseases is not well understood.

Practical challenges drive this lack of understanding. Mitochondrial function is difficult to assay in a high throughput manner. Therefore, most studies use cellular mtDNA content, which can be estimated from sequence data, as a proxy for mitochondrial function, as it can be correlated with the respiratory activity of a cell and mtDNA gene expression [4]. However, associations between mtDNA copy number and common diseases are inconsistent across studies (e.g. see [5] for a review), due to lack of power and differences in tissue type used to estimate mtDNA copy number. Hägg *et al*. is the only well-powered phenome-wide association study (PheWAS) of mtDNA copy number [6]. However, because that study was performed in the UK Biobank, it is not clear whether or not it can be generalized to more diverse cohorts.

In this study, we analyzed genetic and electronic health record data from the Penn Medicine BioBank (PMBB), a large, diverse cohort of African and European ancestry to study the extent to which we can understand the biology underlying genetic and phenotypic correlates of mtDNA copy number. We carried out genome-wide and phenome-wide association studies (GWAS and PheWAS, respectively), separately in two sub-cohorts with largely African (N = 8,598) and European (N = 22,068) ancestry. This allowed us not only to replicate our findings but compare and contrast the genetic basis and phenotypic correlates of mtDNA copy number as a function of ancestry.

## Results

### Peripheral blood mtDNA copy number is a function of cell composition

We estimated mtDNA copy number using the exome sequence data (derived from whole blood) of participants from the Penn Medicine BioBank (PMBB), which has so far recruited more than 175,000 patients with electronic consent through the University of Pennsylvania Health System. We analyzed data from 30,666 unrelated individuals, 22,068 individuals with European ancestry (broadly defined) and 8,598 individuals with mixed African and European ancestry, which we refer to as EUR and AFR cohorts, respectively (analyzed separately). We used the ratio of the mean sequencing depth of off-target reads mapping to the mtDNA to that of reads mapping to an equal number (16,569) of randomly sampled autosomal positions to estimate the average number of mtDNA per cell in whole blood. Note, that because exome sequence data is enriched for autosomal reads relative to mtDNA reads, our estimate does not represent the absolute mtDNA content per cell. Instead, we and other studies that rely on exome sequence or array data, capture the *relative* number of mtDNA copies per cell (which we refer to as rmtCN).

Inter-individual variation in the mtDNA content of whole blood is a function of blood cell composition. We find that the log of rmtCN is largely explained by neutrophil and platelet counts and, to a lesser extent, of other cell types (Fig. 1) in agreement with previous reports [6–8]. The effect of cell composition is consistent in direction between the two cohorts, with neutrophil counts having a negative effect and platelets having a positive effect on rmtCN (Fig. 1). The effect of neutrophil count was larger in the AFR cohort compared to the EUR cohort (Fig. 1) and this difference is not driven by a confounding effect of ancestry, which is associated with neutrophil counts and mtDNA copy number in opposite directions. One possible explanation for this is that neutrophils in the AFR cohort carry fewer mtDNA copies per cell compared to the EUR cohort.

**Figure 1:**
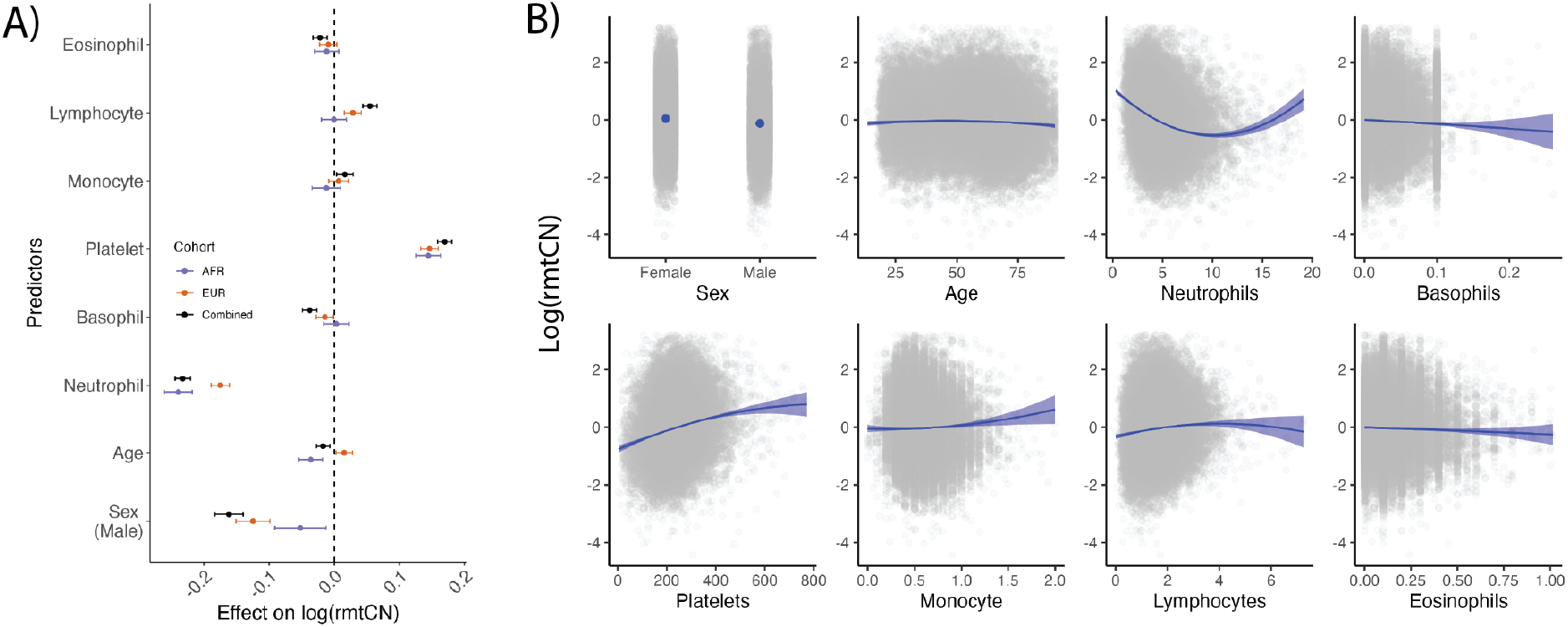
Effect size of sex, age, and blood counts on relative mtDNA copy number (rmtCN). (A) Effect sizes estimated in the combined sample (AFR+EUR) or separately in the AFR and EUR cohorts. The whiskers represent the 95% confidence interval of the point estimate. Effect sizes are displayed in units of standard deviation of lrmtCN. (B) Blue curves represent the predicted values of lrmtCN based on the conditional effects of each predictor (x-axis). Actual data are overlaid as grey points. Age is expressed in years whereas blood counts are expressed in 1000 cells/μL of blood.

The effect of cell type composition was also nonlinear (Fig. 1) and this needs to be appropriately modeled to ensure that downstream analyses are not driven by variation in cell composition. We modeled the log of rmtCN as a function of sex, age, age^2^, and linear and quadratic terms for blood counts (neutrophils, basophils, eosinophils, lymphocytes, monocytes, and platelets). We also included interaction terms to allow the effects of blood counts to vary with age and sex (Methods). The residuals from this model capture variation in the mean number of mtDNA copies per cell independent of blood cell composition, and we hereafter refer to them as rlrmtCN (residual log of rmtCN). Note, however, that the residuals are not informative about whether mtDNA copy number varies across all cell types uniformly or because of a single cell type. To validate that our model appropriately accounts for blood cell composition, we tested whether rlrmtCN was associated with the Duffy-null allele in the AFR cohort. The Duffy-null allele, because it protects red blood cells from infection by *Plasmodium vivax*, is almost fixed in Africa, while being virtually absent elsewhere [9]. The allele is also one of the strongest known associations for neutropenia (low neutrophil count) [10] and thus is expected to be associated with mtDNA copy number if it captures variation in blood cell composition. We confirm this by showing that the Duffy-null allele is significantly associated with the log of rmtCN (*p* = 1.70 *×* 10^−33^) but not with rlrmtCN (*p* = 4.9 *×* 10^−6^) at the genome-wide significance threshold (*p <* 5 *×* 10^−8^) (Fig. S1), suggesting that our model appropriately accounts for at least neutrophil composition. We address any further role of the Duffy-null allele in contributing to mtDNA copy number variation in a later section.

### MtDNA copy number is associated with health-related traits

Because it is correlated with the metabolic activity of the cell, mtDNA copy number is often used as a proxy for mitochondrial (dys)function [5], and changes in mtDNA copy number are a common symptom and sometimes a cause (e.g. mtDNA depletion syndrome) of mitochondrial diseases [11]. To understand if mtDNA copy number is associated with common diseases, we carried out a PheWAS in the PMBB by testing for associations between rlrmtCN and a range of health-related phenotypes (1,353 in the EUR cohort and 1,157 in the AFR cohort), correcting for sex, age, age^2^, and 20 genetic principal components (PCs) as covariates, separately within each cohort.

MtDNA copy number was associated with many diseases related to metabolically active tissues such as liver, heart, and brain that are common targets of mitochondrial dysfunction [12–14]. For example, in the EUR cohort, rlrmtCN was negatively associated with liver damage (7 phecodes at FDR 0.005 and 9 phecodes at FDR 0.05; e.g. liver abscess, cirrhosis, portal hypertension, and esophageal bleeding, and alcoholism; Fig. 2, Table S1). RlrmtCN was also correlated with aspartate aminotransferase (AST) and total bilirubin in the blood, elevated levels of which are both an indicator of alcohol use and alcohol-related liver damage [15] (Fig. 2, Table S1). While none of these associations were significant at the 0.005 FDR in the AFR cohort, their effects were in the same direction (8/8 phenotypes, binomial p-value = 0.008), were correlated (*r*_*beta*_ = 0.65), and were directionally consistent with previously reported association by Hägg *et al*. with esophageal bleeding and portal hypertension [6]. The association between rlrmtCN and liver damage is attenuated, but does not disappear, if we include case/control status for alcoholism as a covariate (Table S2). This suggests that the association between rlrmtCN and liver damage may reflect the causal effcect of alcohol use on both liver damage and rlrmtCN. This is consistent with an experiment in mice showing that an alcohol binge can lead to drastic changes in mtDNA copy number [16].

**Figure 2:**
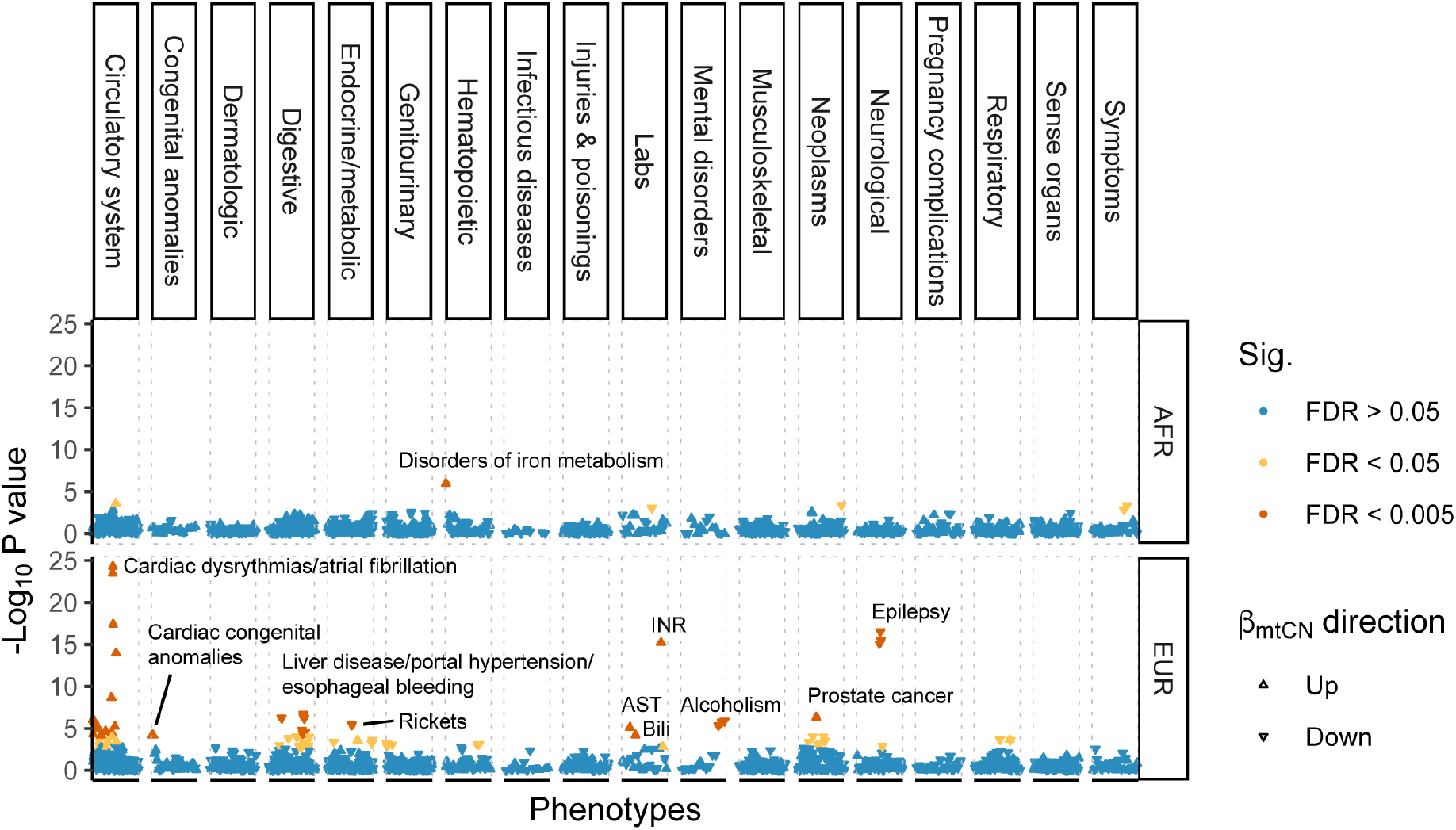
Phenome-wide association study of mtDNA copy number. Phenotypes are ordered on the x-axis and the y-axis shows the -log_10_ p-value of the association with mtDNA copy number separately within the AFR and EUR cohorts. Associations that pass the 0.005 and 0.05 false discovery rate are colored in red and yellow, respectively.

We also observed a positive association between rlrmtCN and cardiac dysfunction — mostly phenotypes related to cardiac dysrhythmias — in the EUR cohort (14 phecodes at FDR 0.005 and 9 phecodes at FDR 0.05; e.g. atrial fibrillation, palpitations, atrial flutter, cardiomyopathy, and mitral valve disease, Fig. 2, Table S1). The associations between rlrmtCN and cardiac phecodes were directionally consistent between the AFR and EUR cohorts (14/14 phecodes, binomial p-value = 1.2 *×* 10^−4^) and were correlated (*r*_*beta*_ = 0.56). The association with cardiac dysrhythmias is also consistent with previous observations of elevated mtDNA copy number in patients with atrial fibrillation [17, 18]. However, our associations are in the opposite direction of the negative association with cardiomegaly reported by Hägg *et al*. [6] and with general cardiovascular disease reported by Ashar *et al*. [19]. We believe that this discrepancy might be explained by differences across studies in how blood cell composition is modeled. Ashar *et al*. [19] do not fully account for blood cell composition or ancestry and Hägg *et al*. only correct for percentage of neutrophils and lymphocytes and total white blood cell count. As an example, we show that the association between mtDNA copy number and some cardiovascular phenotypes are highly sensitive to how blood cell composition is modeled (Fig. S2), suggesting that some previous associations might be driven by blood cell counts as opposed to mtCN *per se*. The association of rlrmtCN with cardiac dysrhythmia phenotypes (e.g. atrial fibrillation) were less sensitive to blood cell composition as they were positively associated with mtDNA copy number in all models (Fig. S2). Altogether, this suggests that mtCN might indeed be a biomarker of some cardiovascular diseases.

The association between some phenotypes and mtDNA copy number was less consistent between the AFR and EUR cohorts. RlrmtCN was negatively associated with epilepsy (3 phecodes at FDR 0.005 and 1 at FDR 0.05), which is a common symptom across mitochondrial disorders, including mtDNA depletion syndrome [20]. But it was not significant in the AFR cohort. In addition to diseases of metabolically active tissues, rlrmtCN was positively associated with prostate cancer and international normalized ratio (INR), which measures the time it takes for blood to clot, and negatively associated with rickets in the EUR cohort (0.005 FDR). In the AFR cohort, rlrmtCN was only associated (positively at 0.005 FDR) with iron metabolism disorders.

### Ancestry-related differences in the heritability of mtDNA copy number

To study the genetic architecture of mtDNA copy number, we first estimated the SNP heritability 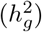 of rlrmtCN in the AFR and EUR cohorts using GCTA [21] with 20 genetic PCs as fixed covariates (Methods). The 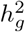 of rlrmtCN in the EUR cohort was 0.10 (95% CI: 0.05 - 0.14), which overlaps with previous estimates (Hägg *et al*. = 0.08, Longchamps *et al*. = 0.07) [6, 22]. The 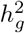 in the AFR cohort was significantly higher at 0.30 (95% CI: 0.20 - 0.39) and we explored a number of explanations for this difference.

First, we suspected that the difference in 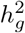 might be driven by known highly differentiated alleles such as the Duffy-null allele, which was not associated with rlrmtCN on a genome-wide level (Fig. S1) but may still contribute to rlrmtCN heritability through its effect on neutrophil counts if the counts were measured imperfectly and/or if they were not correlated with the underlying genetic value. This hypothesis was motivated by the observation that neutrophil heritability is also higher in the AFR cohort (Fig. S3) and a disproportionately large fraction of rlrmtCN 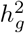 is contributed by chromosome 1 which contains the Duffy locus (Fig. 3). However, including the genotype at the Duffy-null allele (rs2814778), which explains most of the heritability in neutrophil counts in the AFR cohort (Fig. S4), as a fixed effect in the model does not affect rlrmtCN 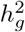 (Fig. 3). Including the genotype for rs73885319 (variant at the *APOL1* locus)—another highly differentiated allele which explains much of the difference in risk of kidney disease between individuals of African and European ancestry [23, 24]—also did not change 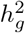 (Fig. S5). This suggests that the difference in 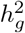 between the AFR and EUR cohorts cannot be explained by these large effect and highly differentiated alleles.

**Figure 3:**
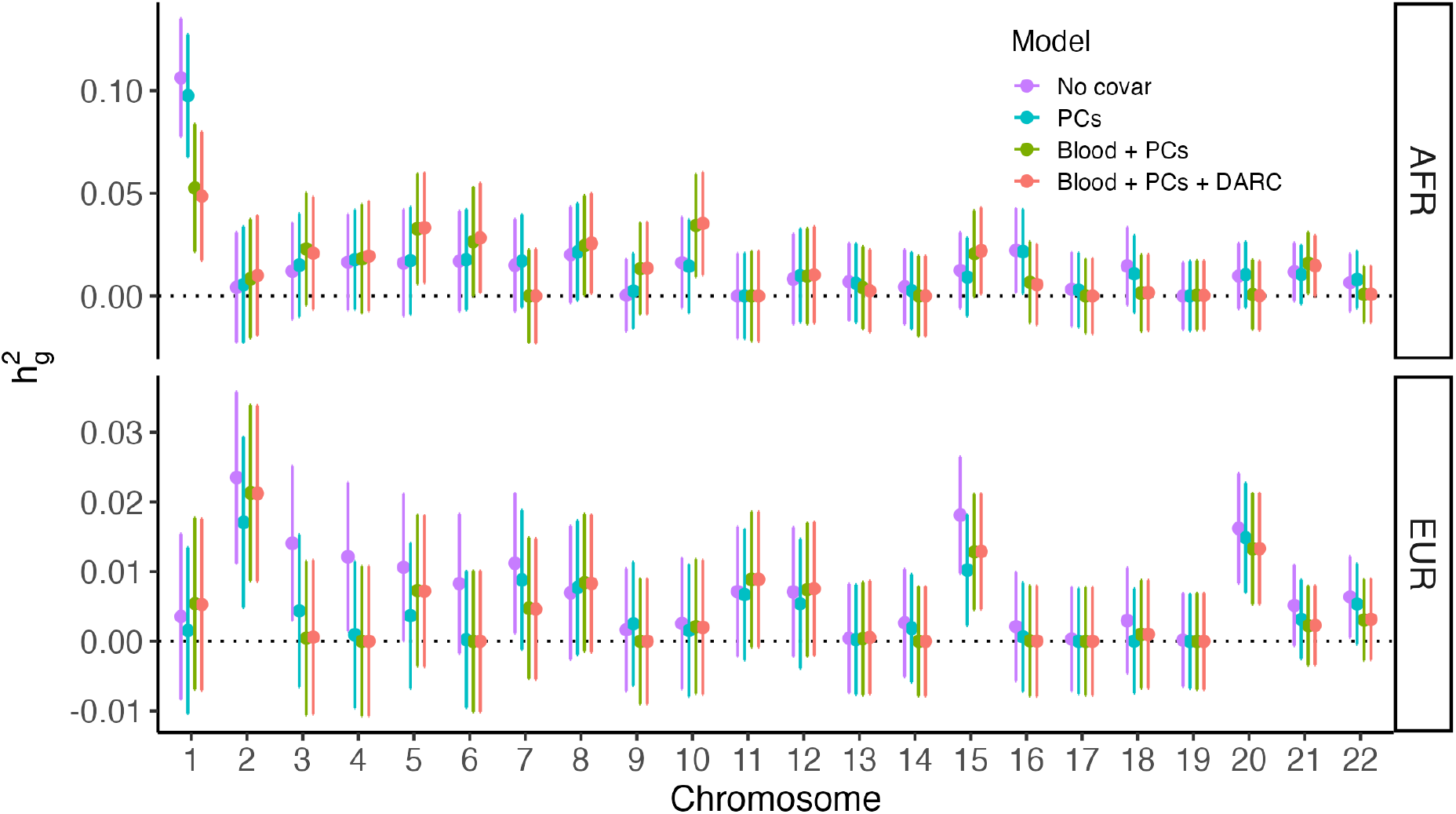
SNP heritability of mtDNA copy number (y-axis) contributed by each chromosome (x-axis). The points represent point estimates and the bars represent the 95% confidence intervals. The colors represent different sets of covariates. No covar = no correction for blood composition, sex, age, or PCs; PCs = correction for sex, age, and PCs; PCs + blood = additional correction for blood cell composition; PCs + blood + DARC = additional correction for Duffy-null genotype.

Second, we asked if rlrmtCN 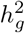 in the AFR cohort could be explained by the heritability underlying blood traits that were not modeled in our analysis. We hypothesized that AST level, which is correlated with rlrmtCN (Fig. 2) and also has a higher heritability in the AFR cohort (Fig. S3), could be contributing to rlrmtCN 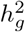 in the AFR cohort. However, including AST level as a fixed covariate in our model did not affect 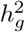 estimates (Fig. S5), suggesting that this is not the explanation for increased heritability in the AFR cohort.

Third, we investigated whether heritability underlying blood traits that were not measured in the PMBB could be driving 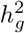 in the AFR cohort. To test this, we constructed polygenic risk scores (PRS) for 15 blood traits using effect sizes estimated in a GWAS carried out in ≈ 500,000 individuals [25] (Methods). We validated these scores by showing that the effect sizes of GWAS variants for blood traits that were measured in the PMBB are correlated with their effects in the EUR cohort (Fig. S6) and that the PRS is correlated with the actual phenotype in both cohorts (Fig. S7). However, including these PRS as covariates also did not affect 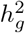 estimates (Fig. S5). We draw two conclusions from this result: First, that unmeasured blood traits are unlikely to contribute to the difference in rlrmtCN heritability between the AFR and EUR cohorts. Second, the EUR-AFR difference in rlrmtCN 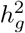 cannot be explained by measurement noise in blood counts in the PMBB.

Finally, we carried out admixture mapping to identify differentiated alleles which might contribute to rlrmtCN in the AFR cohort. To do this, we tested the association between local ancestry across the genome and rlrmtCN in the AFR cohort with the genome-wide ancestry fraction as a covariate. While we did not discover any loci at the genome-wide level, we show that including the genotypes at the most significant hits (21 independent hits at p-value < 0.05, Methods) as covariates in the model substantially reduces rlrmtCN 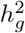 in the AFR cohort to 0.15 (95% CI: 0.05 - 0.14), which overlaps with the 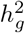 estimatein the EUR cohort. This suggests that the heritability of rlrmtCN in the AFR cohort might be driven by alleles with large frequency differences between individuals of African and European ancestry. Whether these alleles are associated with mtDNA copy number directly or through their effects on other blood traits will require further investigation.

### Similar effects of mtDNA copy number-associated variants in AFR and EUR cohorts

To discover these alleles, we carried out a GWAS of rlrmtCN. We used imputed data and included the first 20 PCs, computed separately in the AFR and EUR cohorts, to correct for population structure. We did not discover any associations at a genome-wide significance threshold of 5 *×* 10^−08^ (Fig. S9) in either cohort. This includes *TFAM*, which was first identified in a smaller sample of ≈ 10K individuals [26]. We then tested if we could replicate other associations discovered previously in much larger GWAS [6, 22, 27]. These studies were largely based on the same dataset (i.e. the UK Biobank) so we restricted our analysis to variants identified in Longchamps *et al*., which was the largest study in terms of sample size [22]. Of the 129 independent variants reported in [22], 110 were present in our imputed data. Of these, only three were significant at a replication threshold of 4.5 *×* 10^−04^ (0.05/110) in the EUR cohort: rs3110823 in the gene *STMP1* (*β*_*A allele*_ = 0.056, *p* = 2.24 *×* 10^−06^), rs10419397 near the gene *USHBP1* (*β*_*A allele*_ = 0.047, *p* = 8.69 *×* 10^−07^), and rs12247015 (*β*_*A allele*_ = 0.039, *p* = 1.14 *×* 10^−05^) in the 5’ UTR of *TFAM*. This is fewer than the 6.4 associations that we expected to replicate (we had *>* 80% power to detect 8 loci) based on the effect sizes estimated in [22] (Table S3, Methods). Nevertheless, the effect sizes of the 110 variants were strongly correlated with their effects in the PMBB (Fig. 4, *ρ*_*EUR*_ = 0.64, *ρ*_*AF R*_ = 0.41). Note, however, that the PMBB effect sizes are smaller, on average, than the effects reported in Longchamps *et al*. (Fig. 4), which explains why we replicated fewer variants than expected. To understand the reason for the downward bias in effect sizes, we considered the possibility that our estimate of mtDNA copy number might be noisier compared to that of Longchamps *et al*.. But this explanation is not likely given that the heritability of rlrmtCN in the EUR cohort is similar to previous studies.

**Figure 4:**
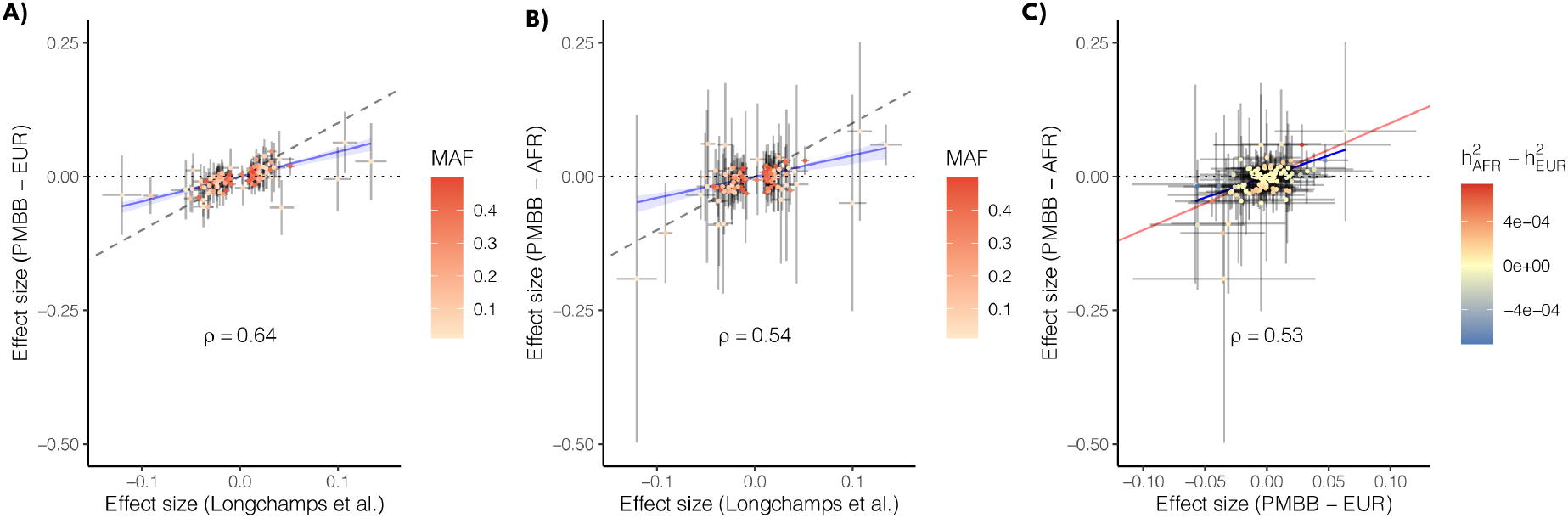
Comparison of effect sizes of mtCN variants discovered in Longchamps *et al*. and their effects re-estimated in the PMBB (A) EUR cohort, and (B) AFR cohort. (C) Comparison of the effects of the same variants between the AFR and EUR cohorts. The color scale in (A) and (B) represents the minor allele frequency in the original study [22] and in (C) represents the difference between the AFR and EUR cohorts in the variance explained by each locus.

The effect sizes of GWAS variants were similar in magnitude and highly correlated between the AFR and EUR cohorts (Fig. 4). The GWAS variants also explain a similar fraction of the phenotypic variance in the two cohorts 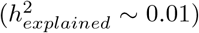. Thus, the difference in heritability between the two cohorts (see previous section) cannot be explained by a difference in the joint distribution of frequency and effect size at GWAS loci.

### No effect of mito-nuclear incompatibility on mtDNA copy number

In a previous study, one of us (A.Z.) found that mtDNA copy number in lymphoblastoid cell lines from admixed individuals was negatively correlated with increasing discordance between the mitochondrial and nuclear genomes such that cells with a higher degree of divergence between nuclear and mitochondrial ancestry exhibited lower mtDNA copy number, on average, than cells where the nuclear and mitochondrial ancestry were similar [28]. This might arise if there were a difference in replication rate between mitochondrial genomes that are more divergent vs similar in ancestry to the individual’s nuclear genome (e.g., due to mito-nuclear incompatibility). We wanted to replicate this result in primary tissue and thus, analyzed data from a subset of individuals from the AFR cohort with mixed African and European ancestry who carried either a European or African haplogroup (N = 8,311, Methods). We fitted a linear model with rlrmtCN as the dependent variable and proportion of African ancestry in the nuclear genome, mtDNA ancestry, and the interaction between mtDNA and nuclear ancestry as predictors. The interaction term was not statistically significant (*β* = −0.31, *p* = 0.095; Fig. 5) contrary to our expectation under the hypothesis that mito-nuclear ancestry discordance leads to a reduction in mtDNA copy number [28]. The discrepancy between the the result shown here and the original study [28] lies in how mito-nuclear discordance is defined. In [28], mito-nuclear discordance was defined as the total fraction of nuclear ancestry that is different in continental original from the mtDNA. For instance, the discordance of someone with the L mtDNA haplogroup (predominantly found in Africa) and 75%, 25%, and 11% of African, Native American, and European ancestry, respectively, in the nuclear genome would be 0.25 + 0.11 = 0.36. This measure has also been used in other studies to test for mito-nuclear incompatibility in admixed individuals [29]. The problem with this measure, however, is that it captures the main effect of nuclear ancestry — which is significantly correlated with mtDNA copy number (Fig. 5) — if mtDNA haplogroups are non-uniformly distributed in the sample (e.g. 80% African and 20% European in the PMBB). As a result, discordance will be associated with copy number, even if there is no effect of incompatibility. We confirm this by showing that mtDNA haplogroup imbalance in the PMBB also causes mito-nuclear discordance to be negatively associated with rlrmtCN (*β* = −0.321, *p* = 3.31 *×* 10^−06^). Therefore, the correct way to test for incompatibility is to test for an interaction between nuclear ancestry and mtDNA haplogroup. Re-analysis of data from [28] shows that that the interaction between mtDNA and nuclear ancestry is also not significantly associated with mtDNA copy number in the original study. Altogether, this shows that there is no evidence for an effect of mito-nuclear incompatibility on mtDNA copy number in admixed individuals.

**Figure 5:**
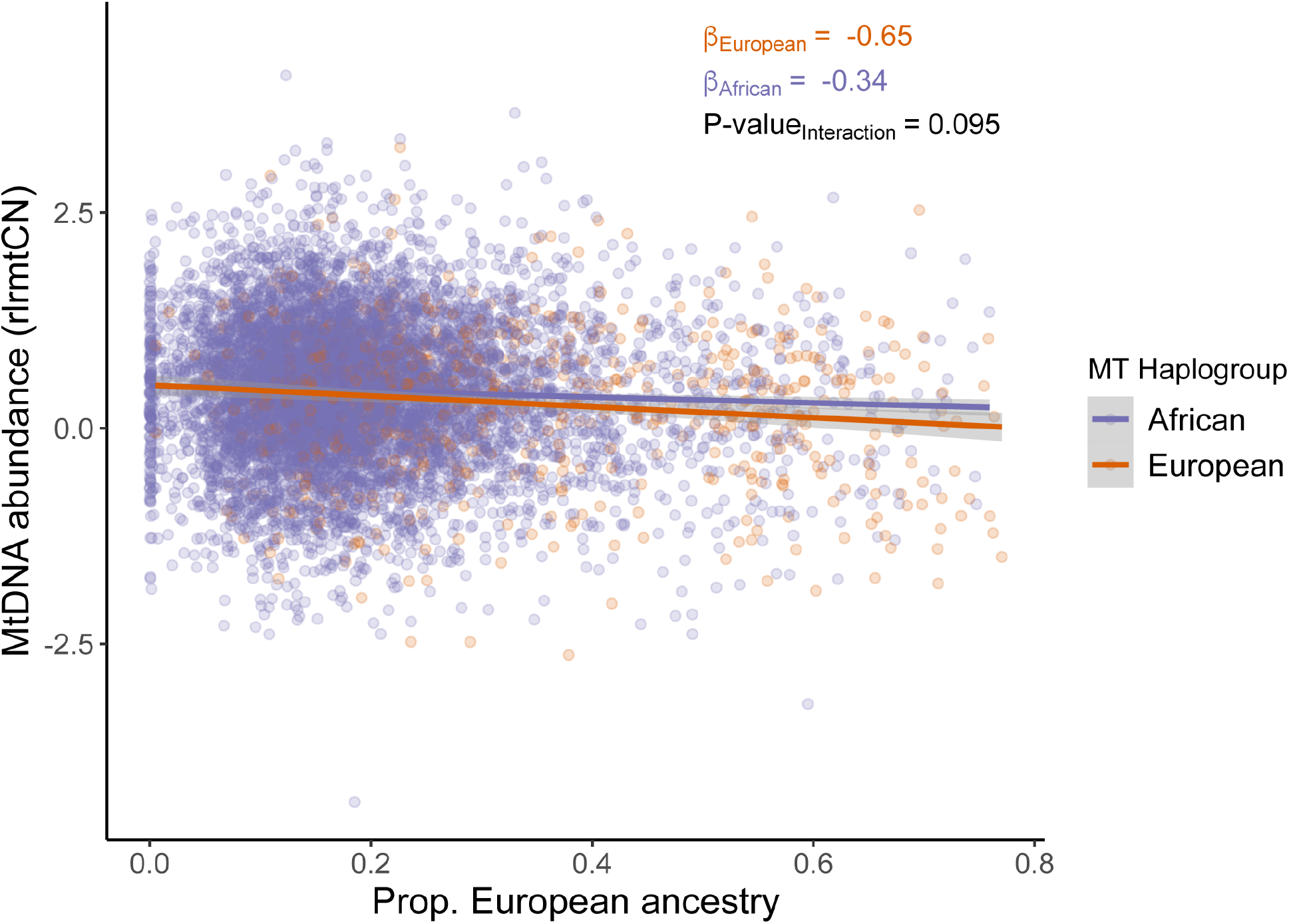
The relationship between nuclear ancestry (x-axis) on residual mtDNA copy number (y-axis) is similar in different mtDNA backgrounds (colors). Variation in mtDNA copy number due to sex, age, age^2^, and blood counts was removed.

**Figure 6:**
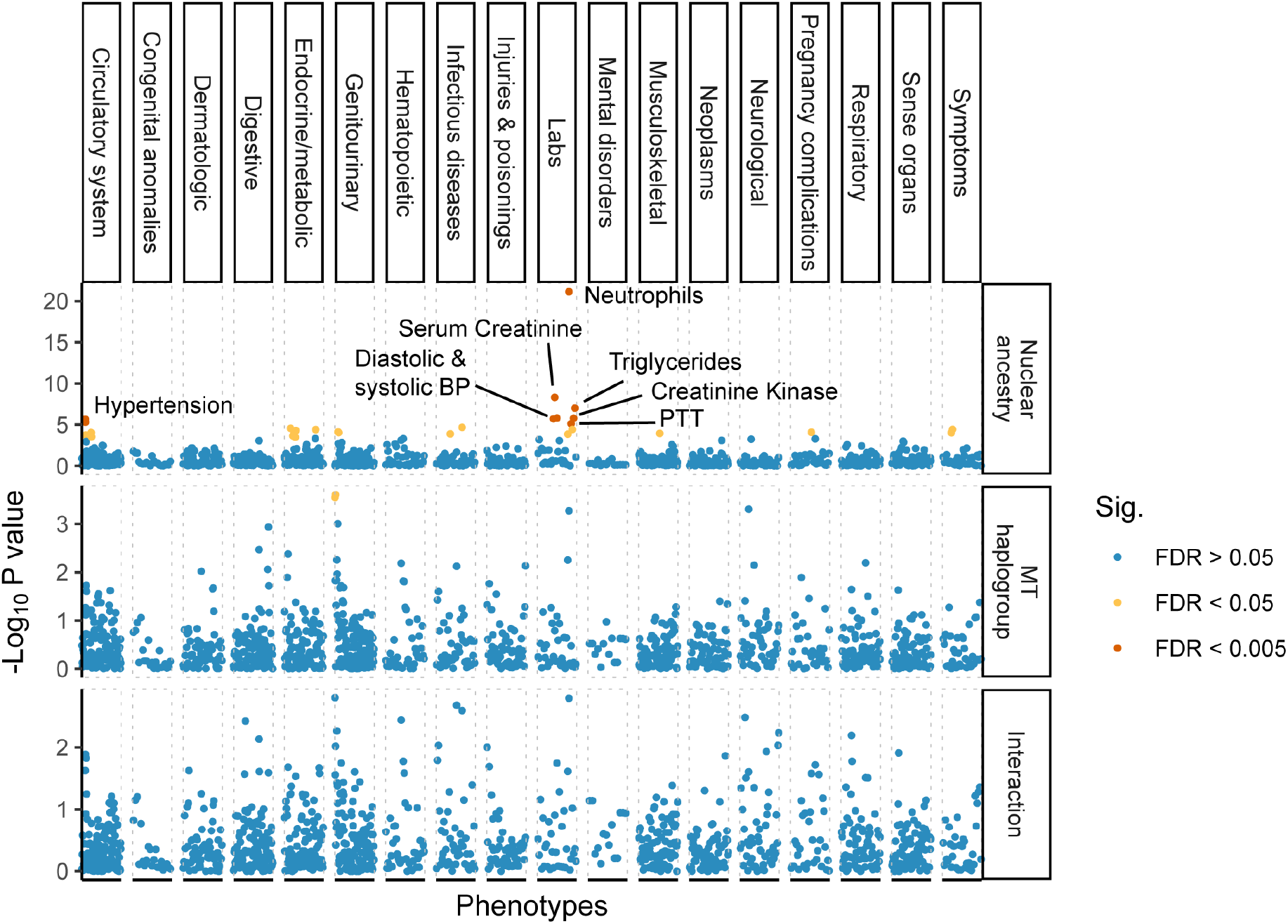
Phenome-wide association of nuclear ancestry (upper), mtDNA haplogrup (middle), and interaction between nuclear and mitochondrial ancestry (lower) in the AFR cohort. Phenotypes are ordered on the x-axis grouped into broader categories based on the PheWAS catalog. Association models were either logistic regression (binary phecodes) or linear (lab measurements) with the same covariates: sex, age, and age^2^. Associations that pass a false discovery rate of 0.005 are highlighted in red while those that pass a FDR of 0.05 are shown in yellow.

### No effect of mito-nuclear incompatibility on health-related traits

We further tested whether there are general phenotypic effects of mito-nuclear incompatibility with a PheWAS on 1,208 health-related phenotypes in the admixed AFR cohort. As before, we fitted ordinary least squares regression for quantitative traits and logistic regression for binary traits, using nuclear ancestry, mtDNA haplogroup, and the interaction between the two as predictors and sex, age, and age^2^ as covariates. The proportion of African ancestry in the nuclear genome was positively correlated (at the 0.005 FDR) with a range of phenotypes including hypertension, hepatitis B, blood pressure (systolic and diastolic), triglyceride levels, serum creatinine levels, and creatine kinase levels, and negatively correlated with neutrophil count — all consistent with worse health outcomes for people with higher African ancestry and also consistent with previously known associations [10, 30, 31]. In fact, of the 1,158 phecodes tested (out of total 1,208 phenotypes), 713 were positively associated with African ancestry (binomial test *p* = 3.25 *×* 10^−15^). In contrast, mtDNA haplogroup was only associated with kidney disease (glomerulonephritis, renal sclerosis) at the 0.05 FDR, with the European haplogroup conferring a higher risk. However, the interaction between mtDNA and nuclear ancestry was not significant at the 0.005 or 0.05 FDR for any phenotype. We show using simulations that, for quantitative traits, we have more than 80% power to detect a negative interaction if the effect of ancestry is larger than 0.5 (in units of standard deviation) (Fig. S10). We have relatively limited power for binary traits, but can detect a negative interaction with 80% probability for common diseases (i.e. prevalence > 0.35) where the effect of ancestry is greater than an odds ratio of 3.5 (Fig. S10). For comparison, the effect of African ancestry on hypertension, which was one of the only binary traits to be significantly associated at the 0.005 FDR, is 1.23, which translates to an odds ratio of 3.4. This suggests that the effects of mito-nuclear incompatibility, if present, are unlikely to be large.

## Discussion

The mitochondrial content (total mitochondrial number and volume) of a cell varies across cell types and is correlated with its mitochondrial activity and bioenergetic needs [4]. Mitochondrial content and activity are also correlated with the number of mtDNA copies in a cell [4], which is easier to assay in a high throughput manner. As such, there has been interest in using mtDNA copy number as a proxy for mitochondrial (dys)function in large scale studies. Many studies have tested for the associations between mtDNA copy number and common diseases (e.g. see [5] for review) but such associations are inconsistent likely because most studies are under-powered and/or they analyze mtDNA copy number from peripheral blood without appropriately accounting for variation in blood cell composition. This makes it difficult to interpret genetic and phenotypic associations of mtDNA copy number. In this study, we analyzed lab measurements and disease outcomes derived from electronic health records, and genetic data to study the genetics of mtDNA copy number and understand the extent to which it is a useful biomarker of health-related phenotypes in a diverse cohort with African and European ancestry.

MtDNA copy number was associated with several health-related phenotypes, particularly those involving metabolically active tissues such as heart and liver. These associations were largely consistent between the AFR and EUR cohorts. For example, there was a negative association with markers of liver damage and a positive association with phenotypes related to certain cardiovascular diseases. The association between mtDNA copy number and liver damage seems to be mediated largely by alcohol use. The positive association between copy number and atrial fibrillation is consistent with previous studies [17, 18] and is thought to be driven by an increase in cell-free circulating mtDNA released by cardiomyocytes in patients with atrial fibrillation, as a result of mitochondrial dysfunction [18]. The association between copy number and other cardiovascular disease (e.g. cardiomegaly) was in the opposite direction to previous reports [6] and we show that this discrepancy might be due, in part, to differences in how blood cell composition is modeled across studies. Some phenotypic associations were also different within our study between the AFR and EUR cohorts. For example, epilepsy, which is a common symptom of mitochondrial disorders, including mtDNA depletion syndrome, was negatively correlated with mtDNA copy number but only in the EUR cohort. We suspect that these discrepancies might be driven by differences in environment (e.g. alcohol use) that also vary with ancestry [32, 33]. Altogether, our results suggest that mtDNA copy number is associated with a range of diseases, but most of these associations are difficult to interpret because they are sensitive to environmental and cellular heterogeneity in peripheral blood. That said, some associations (e.g. alcohol-related liver disease and atrial fibrillation) were robust and replicated across ancestry groups, suggesting that mtDNA copy number might be a useful biomarker of some diseases.

The genetic architecture of mtDNA copy number was less sensitive to blood cell composition and other modifiers, but was strongly associated with ancestry. The heritability of mtDNA copy number was higher in the AFR cohort (∼ 30%) compared to the EUR cohort (∼ 10%). This difference was not driven by the heritability of blood traits that vary with ancestry (e.g. neutrophil counts and AST levels). In fact, we found that the effect sizes of variants discovered in previous GWAS were highly correlated with their effects in our study, in both EUR and AFR cohorts, despite differences in phenotype construction and ancestry. Interestingly, the difference in heritability of mtDNA copy number between the AFR and EUR cohort was not due to a difference in the frequency or effect sizes of associated variants discovered in previous GWAS [22]. Instead, our admixture mapping analysis suggests that the difference in heritability between the AFR and EUR cohorts might be driven by variants that are common in AFR, but rare in European ancestry populations, and were therefore not discovered in previous GWAS. If true, larger multi-ethnic studies would be needed to discover these variants and to understand whether they affect mtDNA copy number directly or through other phenotypes that are more heritable among individuals of African ancestry.

We also tested for an effect of mito-nuclear incompatibility on mtDNA copy number. Mito-nuclear incompatibilities have been demonstrated in other organisms (e.g. drosophila and marine copepods [34– 36]) but the extent to which they contribute to human disease risk is debated [37, 38]. An analysis of cell lines from admixed individuals from the 1000 Genomes Project previously showed that increasing mito-nuclear ancestry discordance, measured as the fraction of autosomal ancestry that is divergent from mtDNA ancestry leads to a reduction in mtDNA copy number decreases [28]. This was interpreted to be an effect of incompatibility between mitochondrial and nuclear ancestry. However, here we show that this does not replicate in primary tissue from a much larger sample of admixed individuals and further show that the original result was due to a statistical artifact that captured the effect of nuclear ancestry, as opposed to that of incompatibility. In conclusion, there is so far no evidence that mito-nuclear incompatibility affects mtDNA copy number in admixed individuals. We also did not detect any significant effects of mito-nuclear incompatibility in a phenome-wide analysis of 1,208 health-related phenotypes. For example, we did not observe an effect of mito-nuclear ancestry interactions on any pregnancy-related phenotypes (e.g. miscarriage, stillbirth, early onset delivery, pre-term birth, and preeclampsia) in contrast to a previous study [29]. That we did not detect such effects in admixed individuals suggests that they do not contribute substantially to variation in medically-relevant phenotypes.

A limitation of our study is that phenotypic associations of mtDNA copy number seem to be mediated by and are sensitive to genetic and environmental modifiers (e.g. ancestry, blood cell composition, and alcohol use). Differences between studies in the distribution of such modifiers and how they are modeled can lead to different results. Blood cell composition can also vary quite drastically with time [39] and complete blood counts in studies such as the PMBB and UK Biobank may not come from the same samples that were used to estimate mtDNA copy number. Thus, even the best methods of correction for blood composition cannot guarantee that the associations that we and others observe are independent of the effect of blood counts. As such, the utility of peripheral blood as a reliable biomarker of mitochondrial dysfunction seems, at present, limited. These limitations are not unique to mtDNA copy number but to analyses of all cellular readouts (e.g. gene expression) measured in heterogeneous tissues. That said, the availability of deep phenotype, genetic, and environmental data in population-scale studies such as the UK Biobank and PMBB can be useful in mediation analyses, as we illustrate here, to parse out some of the causal mechanisms underlying phenotypic and genetic associations of mtDNA copy number.

## Methods

### Description of the dataset

All individuals were patients of the University of Pennsylvania Health System and were enrolled in the Penn Medicine BioBank. Written consent was obtained to collect and store biological specimens and electronic health record (EHR) data, and carry out DNA extraction and sequencing. Access and analysis of data for this study was approved by the Institutional Review Board at the University of Pennsylvania.

We started with genetic and EHR data from a total of 39,185 unrelated individuals, who were analyzed in two groups: 10,183 individuals with mixed African and European ancestry and 29,002 individuals with European ancestry (defined broadly), hereafter called AFR and EUR cohorts. Laboratory measurements and disease outcomes were derived from patients’ EHR data. Disease outcomes were obtained as ICD-9 and ICD-10 codes, which we mapped to Phecodes [40, 41]. We defined case/control status for each phecode based on the number of hospital visits, classifying an individual as ‘case’ if they presented in the system with the same phecode at least twice and as ‘controls’ if they were not listed with that phecode at all. Individuals listed once were set to missing. We restricted the analysis to phecodes with more than 20 cases in each of the two cohorts, leading to a total of 1,157 phecodes in the AFR cohort and 1,353 phecodes in the EUR cohort. We also analyzed 25 quantitative laboratory measurements, using the median for each individual if they had multiple measurements. A complete list of phenotypes analyzed for each analysis is available in Tables S1 and S4. For lab measurements, we removed outliers which were > 7 standard deviations away from the mean and used Box-Cox power transformations. Of the initial sample, we had non-missing complete blood count data for 30,666 individuals (EUR = 22,068 and AFR = 8,598) who were then retained for all further analyses.

### Calling mtDNA copy number

Mitochondrial DNA copy number represents the number of copies of mtDNA per cell and can be estimated from whole genome sequence data as twice the ratio of mtDNA depth and autosomal depth. Because exome-sequencing involves enrichment of coding sequences in the nuclear genome, we cannot estimate mtDNA copy number in absolute terms (i.e. in number of copies per cell) but can still capture the relative variation in copy number among individuals. To do this, we used *bcftools mpileup* (version 1.12) to call genotypes from reads aligning to the revised Cambridge Reference Sequence (rCRS) of the human mitochondrial genome, filtering out reads with map quality less than 20 (*-m 20*) and base pair quality less than 30 (*-q 30*). Next, we extracted the depth at each position using *bcftools query -f* ‘%POS%[-:DP]’, giving us an overall mean depth of 2.8x per site per individual. We observed a spike in sequencing depth between 2.5 kbp and 3 kbp on the rCRS (Fig. S11), which has been reported previously [42]. We masked out this region when calculating mean mtDNA depth. We calculated mean autosomal depth across 16,569 sites sampled uniformly at random across the exome. Finally, we took the ratio of mean mtDNA sequencing depth and mean autosomal sequencing depth to get relative mtDNA copy number (rmtCN).

We modeled the log of rmtCN as a function of sex, age, and blood composition using the following linear model in the total sample (AFR and EUR combined) in R:

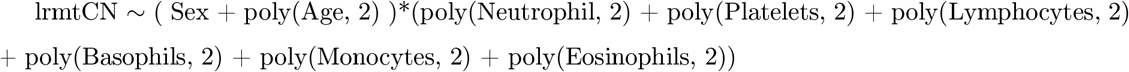

This model accounts for non-linear effects of blood cell counts and allows these effects to vary between males and females and with age. We used the residuals from this model as estimates of mtDNA copy number in all subsequent analyses and referred to them as rlrmtCN.

### mtDNA haplogroup calling and ancestry estimation

We used genotypes at 779 mtDNA SNPs that were genotyped on the GSA array to call haplogroups for each individual with Haplogrep v2.40 using the *classify* function with the *--chip* flag. We validated haplogroup calls by calling haplogroups from exome sequence data using off-target reads aligning to the mitochondrial genome. We show that called haplogroups are highly concordant between exome sequence and snp array data (99% concordance at the top level).

To carry out local ancestry inference, we first phased the genotype data (545,267 SNPs) from the AFR cohort using Beagle version 5.4 [43] and then used RFMix [44] to infer local ancestry (k = 2) with genotypes from the 1000 Genomes Project (CEU and YRI) [45] as reference. We averaged local ancestry for each individual across SNPs which were called with a posterior probability greater than 0.9 to calculate the overall proportion of African and European ancestry. Global ancestry calculated using RFMix was highly correlated (*r*^2^ = 0.99) with unsupervised ancestry estimates generated using ADMIXTURE (k = 2) [46].

### GWAS, heritability, and PheWAS of mtDNA copy number

We carried out GWAS on rlrmtCN against 10,868,495 autosomal markers, which were imputed using the Michigan Imputation Server [47], with the first 20 genetic principal components (PCs) as covariates. PCs were computed separately within each (AFR and EUR) cohort from a genetic relationship matrix (GRM) generated using GCTA version 1.93.2beta [21] from common (MAF > 1%), LD-pruned (*plink --indep-pairwise 100 10 0*.*1* [48]) autosomal SNPs that were directly typed on the array.

We carried out admixture mapping in the AFR cohort by testing the association of rlrmtCN with local ancestry at each variant across the genome using a linear model with the global ancestry proportion and genotype for the Duffy-null allele as covariates. The multiple testing burden in admixture mapping tends to be less than that of GWAS because of long-distance correlations in local ancestry that arise due to admixture. We empirically estimated this testing burden using the approach of Shriner *et al*. [49]. Briefly, we estimated the effective number of tests (*N*_*eff*_) by fitting an autoregressive model to the vector of local ancestry for each chromosome of each individual. This was done using the *effectiveSize()* function in the CODA package in R [50, 51]. We summed this number across chromosomes for each individual and then took the mean across individuals to get *N*_*eff*_, which was 17,821 in our case, resulting in a genome-wide significance threshold of 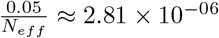.

We used GCTA to estimate the SNP-based heritability of rlrmtCN with the first 20 PCs as fixed effects. We included sex, age, age^2^ in addition to the PCs when estimating 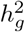 for lab measurements. We included additional covariates (e.g. Duffy-null genotype, see Results section) to determine the source of rlrmtCN heritability in the AFR cohort. The Duffy-null genotype was coded as two variables representing additive (∈ (0, 1, 2)) and dominance effects (0 for homozygotes and 1 for heterozygotes). To determine if the rlrmtCN heritability in the AFR cohort was driven by unknown differentiated alleles, we selected 12 independent loci from the admixture mapping by clumping at a p-value threshold of 0.05 and physical distance of 1Mb and included the genotypes at these loci as fixed effects (in addition to 20 PCs).

PheWAS for rlrmtCN was carried out using a linear model (for quantitative traits) and logistic regression (for binary traits) with age, age^2^, sex, genetic PCs 1-20. We restricted the analysis to phecodes with at least 20 cases. For lab measurements, we removed outliers which were > 7 standard deviations away from the mean and used Box-Cox power transformations where necessary. We used the *boxcox* function in the MASS package in R and estimated *λ* for the residuals of the following model: y ∼ poly(Age, 2) + Sex + 20PCs where y is the quantitative trait of interest.

### Polygenic risk scores

We constructed polygenic risk scores (PRS) for 15 blood traits using the variants discovered in Chen *et al*. [25]. We used the summary statistics from the GWAS carried out in individuals of European ancestry (N ≈ 500,000) available from Table S3 of Chen *et al*. [25]. We retained only SNPs for which the alleles matched between Chen *et al*. and the imputed PMBB genotype data. A comparison between the effect size estimates between Chen *et al*. and this study is provided in Fig. S6. The effect size of one variant (chr1:209451397:G:A) on platelet count as estimated in Chen *et al*. (*β*_*Gallele*_ = −4.4) was much larger than the other variants and in comparison to its estimate in the PMBB (Fig. S6). Their estimate is likely inflated, especially given that the allele is very rare (MAF = <0.001 in their study). We removed this and another rare variant (chr10:122775741:A:G) which had a large effect on mean corpuscular volume of (*β*_*Aallele*_ = −3.49) before calculating PRS. This led to a total of 4,394 variants across all traits, which were used to calculate PRS with the *--score* flag in PLINK [48]. To validate our calculation, we showed that the PRS were correlated with actual values for traits that were available in the PMBB (i.e. neutrophil, monocyte, platelet, lymphocyte, basophil, eosinophil, and white blood cell count) (Fig. S7).

### Power to replicate known associations

To estimate the power to replicate known associations for mtDNA copy number, we downloaded Table S6 from Longchamps *et al*. [22], which contains the list of 129 genome-wide significant SNPs, their positions, and effect sizes. We calculated the power to discover the 110 SNPs that were imputed in the PMBB at the *α* = 4.5 *×* 10^−4^ level of significance (0.05/110 SNPs):

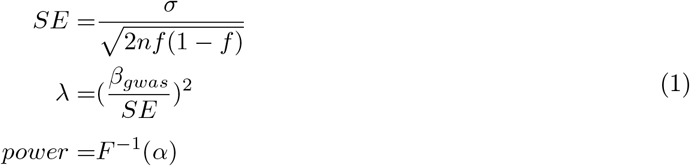

where *σ* ≈ 0.8 is the residual standard deviation in each cohort (AFR and EUR) after accounting for variance due to age, age^2^, sex, and blood cell counts, *f* is the frequency of the effect allele in the cohort, *β*_*gwas*_ is the effect size from the discovery GWAS [22], and *F* is the cumulative distribution function of a chi-square distribution with non-centrality parameter *λ* and one degree of freedom.

We calculated the heritability explained by GWAS variants separately in each cohort (*c*) as 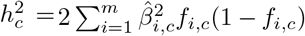 where 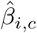 is the effect size estimate of the *i*^*th*^ variant and *f*_*i,c*_ is the minor allele frequency in cohort *c*.

### Analysis of mito-nuclear incompatibility

For the analysis of mito-nuclear incompatibility, we analyzed data from the admixed AFR cohort. We classified haplogroups H, I, J, K, N, R, T, U, V, W, X as “European” and the L haplogroups as “African”. Individuals carrying any other haplogroups (N = 271) were removed, resulting in a total of 8,311 individuals. We fit a logistic regression model (linear if the trait was quantitative) with nuclear ancestry, mtDNA haplogroup, and the interaction between the two as predictors, and sex, age, and age^2^ as covariates. We treated mtDNA haplogroup as a factor with the African haplogroup as the reference level.

We calculated power to test for mito-nuclear incompatibility using simulations. We simulated a quantitative trait with effects of sex, age, age^2^, nuclear ancestry, mtDNA haplogroup, and the interaction between haplogroup and ancestry. We used the effects of sex, age, age^2^ estimated from our data and assumed that the effect of ancestry ranges from 0.05 to 1 (in units of standard deviation of the phenotype). We further assumed a simple model of mito-nuclear incompatibility such that the direction of effect of ancestry is reversed between the two mtDNA haplogroups. We added random noise from a normal distribution with mean zero and standard deviation *σ*, which was also estimated from the data (after removing variation due to covariates) for each trait separately.

For binary (disease status) traits, we selected the effect size of ancestry ranging from an odds ratio of 1.5 to 4. Unlike linear models, the power of the test in a logistic regression depends on the intercept term, which specifies the prevalence of the disease in the population. To model this, we fit a logistic regression model to case status for each binary trait with sex, age, and age^2^ as predictors. Then, we used the estimated coefficients and the mean value of these predictors from the data to generate the intercept: 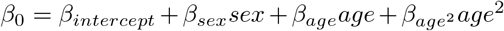. Now let *x*_*j*_ ∈ *{*−1, 1*}* be an indicator variable coding for the mtDNA haplogroup of individual *j, z*_*j*_ ∈ [0, 1] be nuclear ancestry, and *β*_1_ be the (assumed) effect size of ancestry. Then, we can simulate case/control status (*y*_*j*_) for the individual as a bernoulli random variable with probability *π*_*j*_, where:

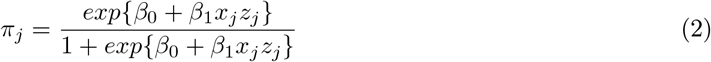

We fitted a logistic regression model (linear regression for quantitative traits) to the simulated data and evaluated significance of the interaction between mitochondrial and nuclear ancestry if the p-value was less than 3.5 *×* 10^−5^ (0.05/1137 traits). We repeated this 1,000 times and calculated power as the fraction of iterations where the interaction term was significant.

## Data Availability

Individual-level genotype and phenotype data from the PMBB are not publicly available due to privacy concerns. However, all summary statistics relevant to this work are made available in Supplementary Tables S1-S4. The code is publicly available and can be accessed on GitHub (https://github.com/Arslan-Zaidi/mtcn\_pmbb).

https://github.com/Arslan-Zaidi/mtcn_pmbb

## Code and data availability

Individual-level genotype and phenotype data from the PMBB are not publicly available due to privacy concerns. However, all summary statistics relevant to this work are made available in Supplementary Tables S1-S4. The code is publicly available and can be accessed on GitHub (https://github.com/ArslanZaidi/mtcn_pmbb).

## Acknowledgements

We thank the patient-participants of Penn Medicine who consented to participate in this research program. We would also like to thank the Penn Medicine BioBank team and Regeneron Genetics Center for providing genetic variant data for analysis. The PMBB is approved under the University of Pennsylvania IRB protocol #813913. The Penn Medicine BioBank is supported by the Perelman School of Medicine at University of Pennsylvania, a gift from the Smilow family, and the National Center for Advancing Translational Sciences of the National Institutes of Health under CTSA award number UL1TR001878. This study was funded by NIGMS awards K99GM137076 (to A.Z.) and R35GM133708 (to I.M.). The content of this paper is solely the responsibility of the authors and does not necessarily represent the official views of the NIH.

## Supplementary material

### Penn Medicine Biobank Team and Contributions

#### Leadership

Daniel J. Rader, M.D., Marylyn D. Ritchie, Ph.D., Michael D. Feldman M.D. Contribution: Contributed to securing funding, study design and oversight.

#### Patient Recruitment and Regulatory Oversight

JoEllen Weaver, Nawar Naseer, Ph.D., M.P.H., Afiya Poindexter, Ashlei Brock, Khadijah Hu-Sain, Yi-An Ko

Contributions: JW manages patient recruitment and regulatory oversight of study. NN manages participant engagement, assists with regulatory oversight, and researcher access. AP, AB, KH, YK perform recruitment and enrollment of study participants.

#### Lab Operations

JoEllen Weaver, Meghan Livingstone, Fred Vadivieso, Ashley Kloter, Stephanie DerOhannessian, Teo Tran, Linda Morrel, Ned Haubein, Joseph Dunn

Contribution: JW, ML, FV, SD conduct oversight of lab operations. ML, FV, AK, SD, TT, LM perform sample processing. NH, JD are responsible for sample tracking and the laboratory information management system.

#### Clinical Informatics

Anurag Verma, Ph.D., Colleen Morse, M.S., Marjorie Risman, M.S., Renae Judy, B.S.

Contribution: All authors contributed to the development and validation of clinical phenotypes used to identify study subjects and (when applicable) controls.

#### Genome Informatics

Anurag Verma Ph.D., Shefali S. Verma, Ph.D., Yuki Bradford, M.S., Scott Dudek, M.S., Theodore Drivas, M.D., PH.D.

Contribution: A.V., S.S.V. are responsible for the analysis, design, and infrastructure needed to quality control genotype and exome data. Y.B. performs the analysis. T.D. and A.V. provides variant and gene annotations and their functional interpretation of variants.

**Figure S1:**
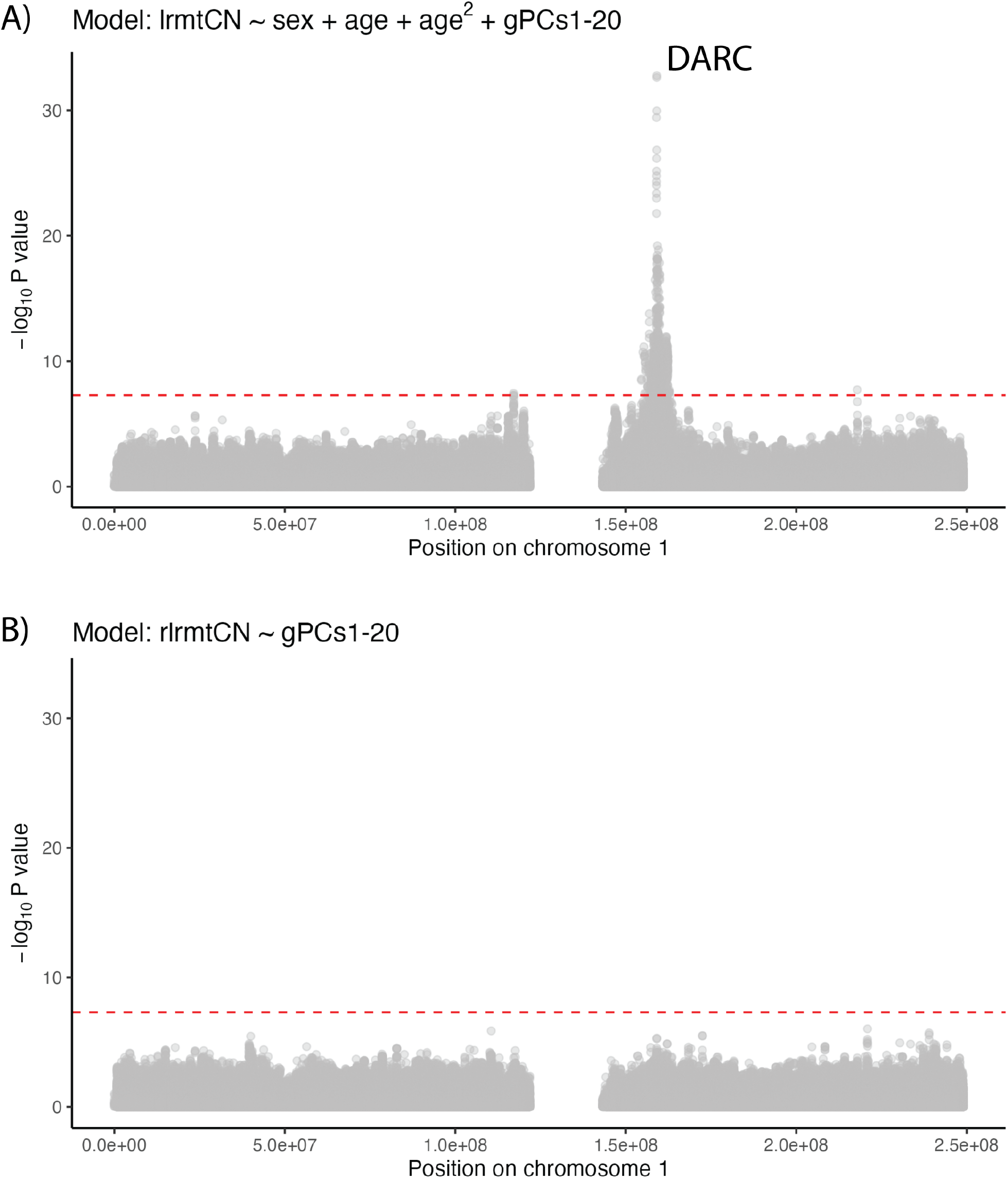
Manhattan plot of GWAS on rmtCN (A) before and (B) after correction for blood cell counts in the AFR cohort. Only chromosome 1 is displayed.

**Figure S2:**
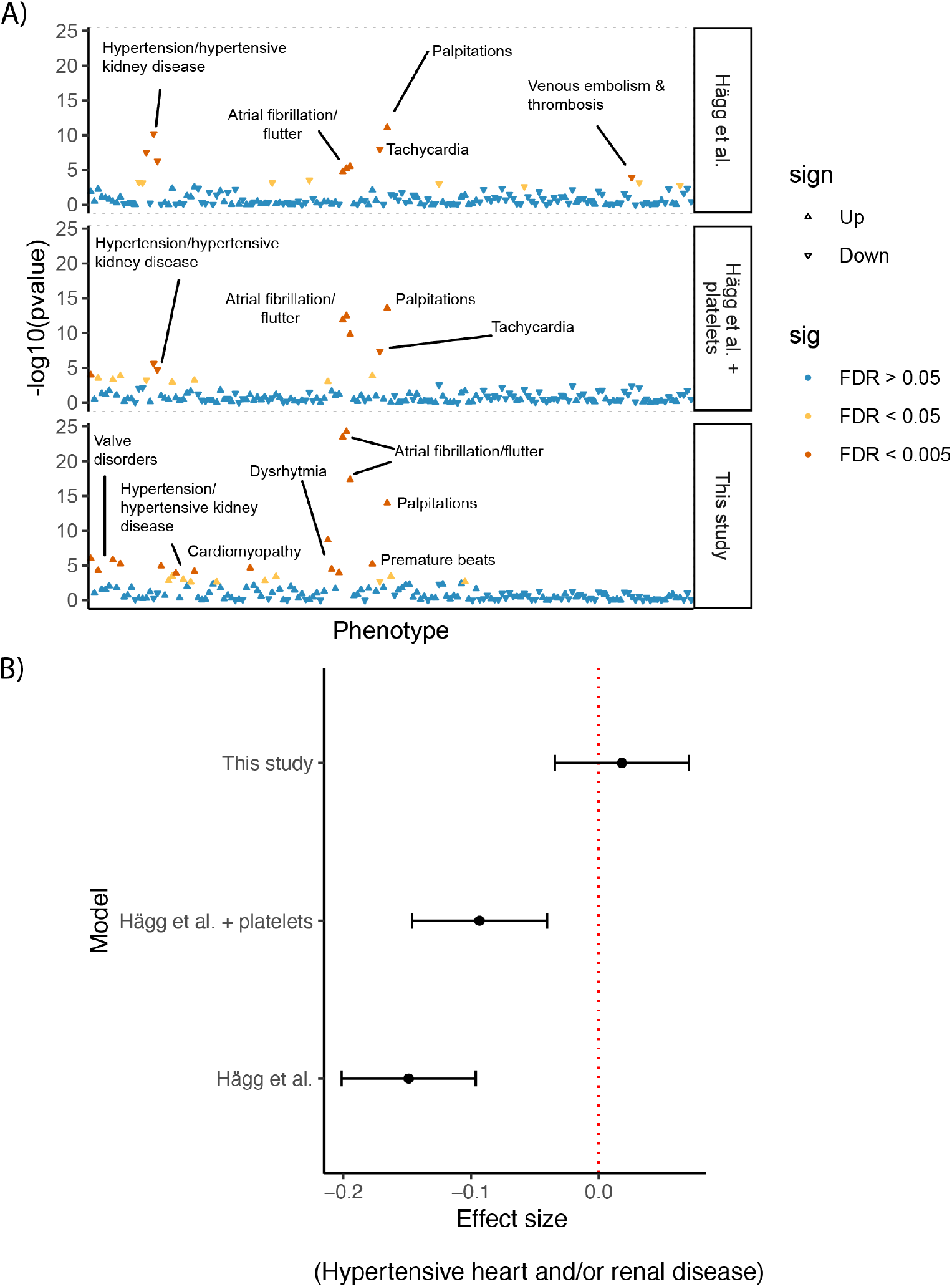
Sensitivity of association of mtDNA copy number with cardiac phenotypes to model choice. (A) The x-axis shows phenotypes arranged in order of phecode number such that similar phenotypes cluster together, and the y-axis shows the negative log of the association p-value. (Top panel) A model closely mimicking that used by Hägg *et al*. where, in addition to lrmtCN, we include sex, age, age^2^, neutrophil %, lymphocyte %, total white blood cell count, and 20 PCs as predictors. (Middel panel) The same as previous model except for the addition of platelet count as a covariate. (Bottom panel) The model used in this study where we use rlrmtCN (residuals from the model described in the main text), sex, age, and age^2^, and 20 PCs as predictors. (B) Forest plot illustrating the change in effect size for one phenotype (hypertension and renal disease) in the EUR cohort.

**Figure S3:**
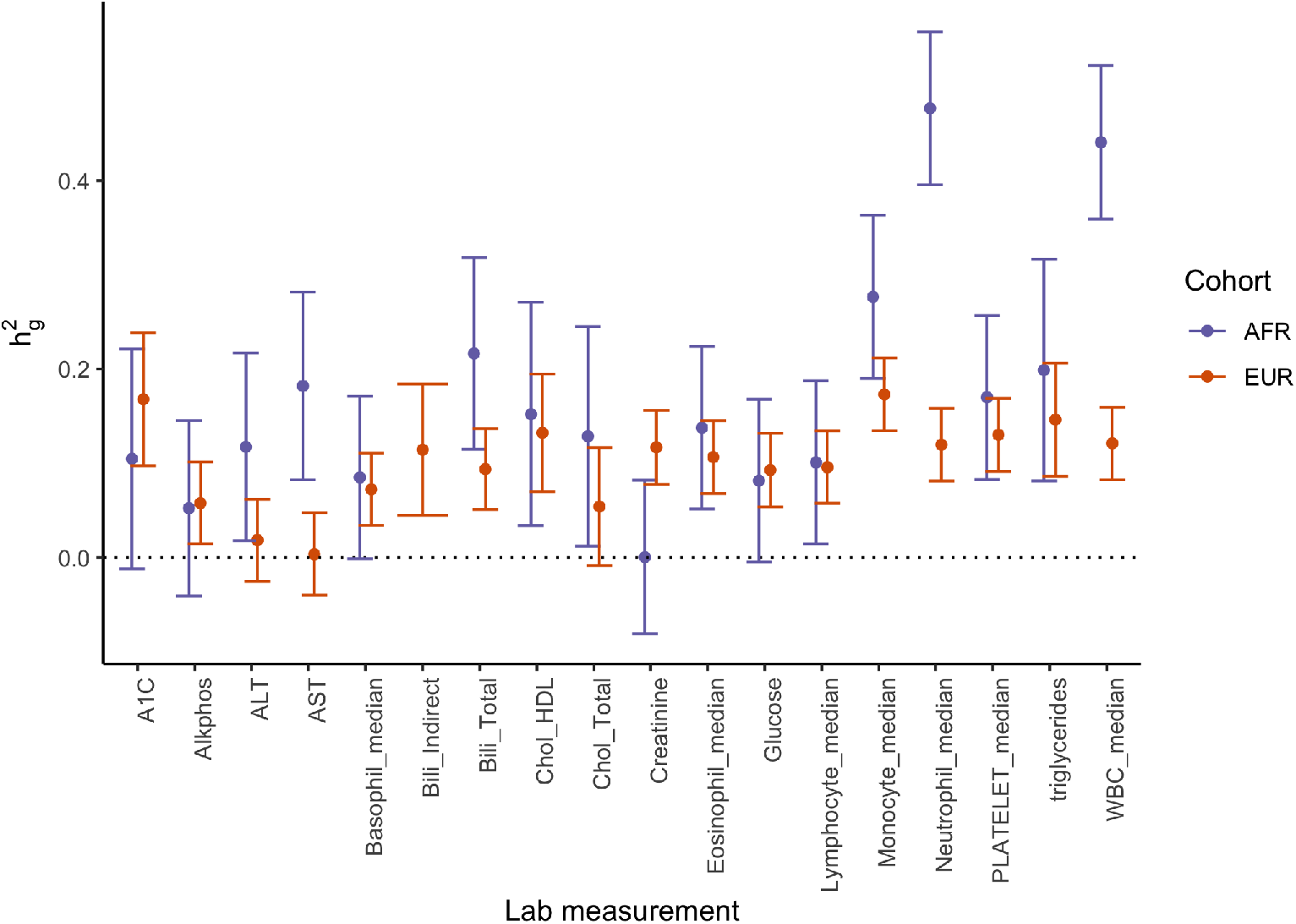
The heritability of lab measurements in the PMBB shown separately for the AFR and EUR cohort. Only lab measurements where the lower bound of the 95% CI was greater than 0 in at least in one of the cohorts is shown

**Figure S4:**
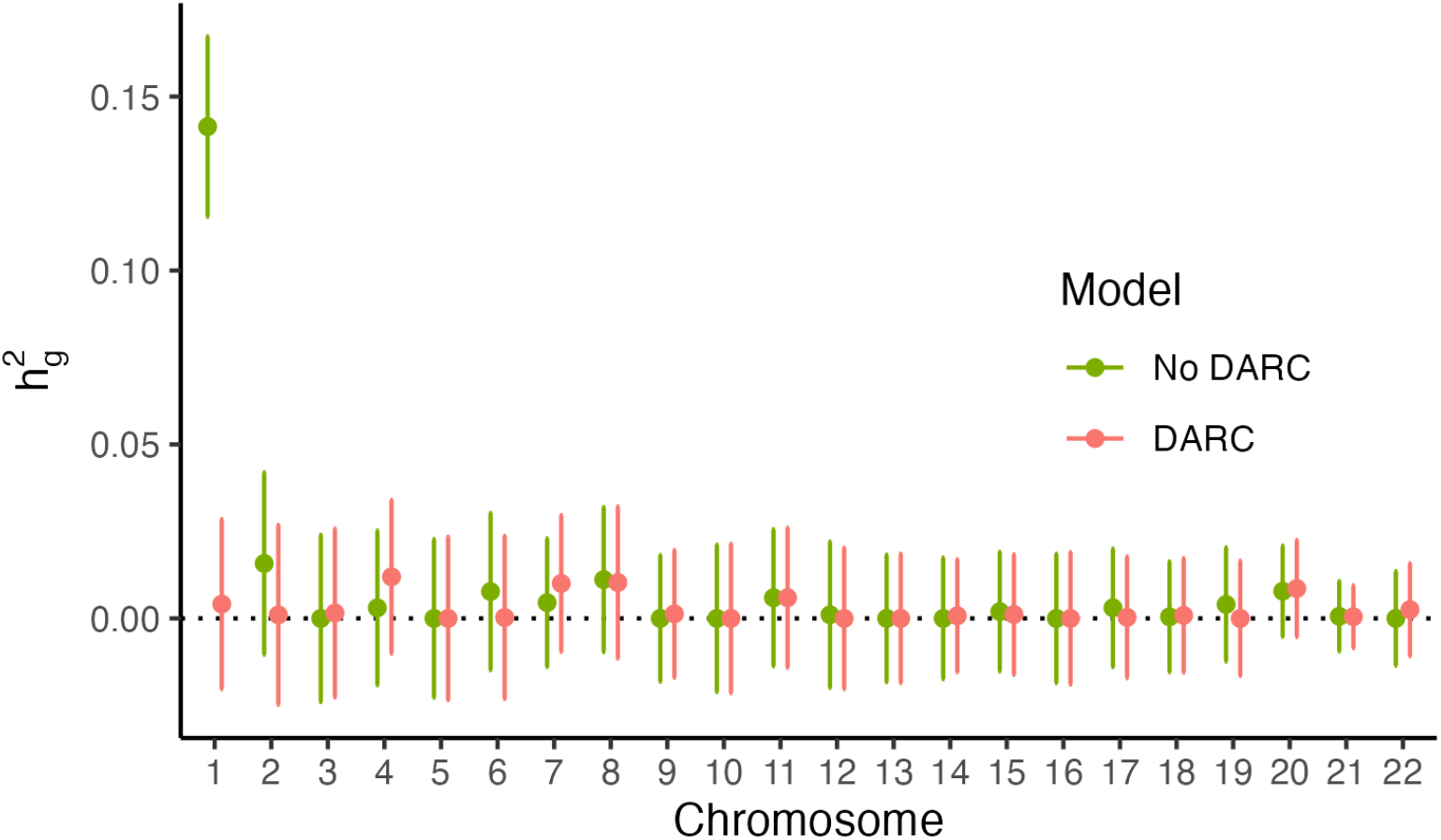
Heritability of neutrophil count partitioned by chromosome in the AFR cohort. The colors represent two models with and without genotype at the Duffy-null allele as a covariate. Both models included sex, age, age^2^, and 20 PCs as covariates.

**Figure S5:**
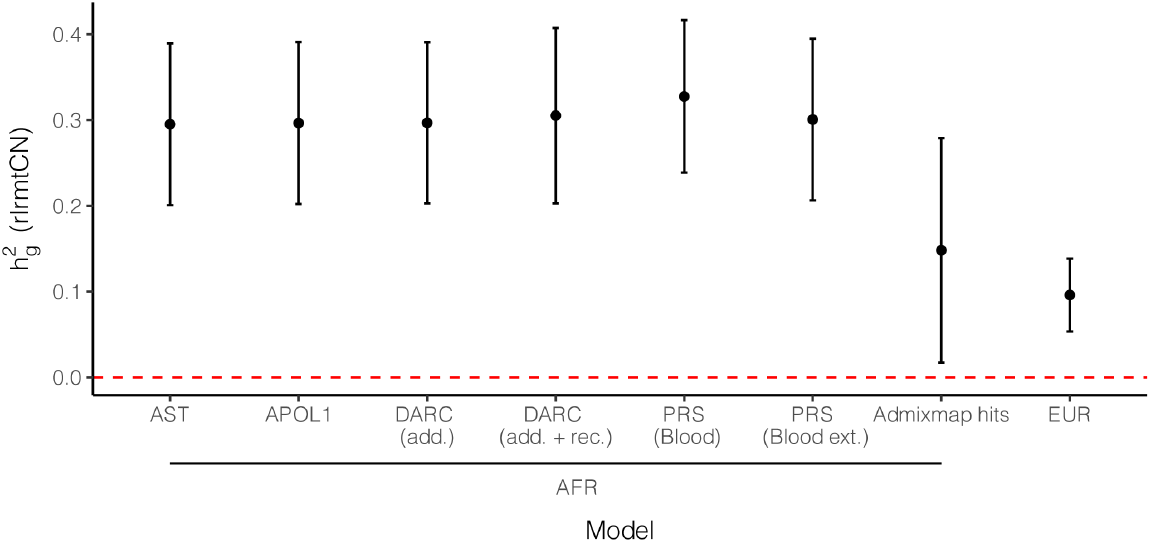
SNP heritability of rlrmtCN in the AFR (first 7 columns) and EUR (last columns) cohorts estimated using GCTA. All models included 20 genetic PCs calculated separately in each cohort. For the AFR cohort, the heritability was estimated with additional covariates: AST = amino aspartate transferase levels; APOL1 = genotype at the APOL1 locus; DARC (add.) = genotype at the rs2814778 SNP coded additively; DARC (add. + rec.) = additive and recessive coding for rs2814778; PRS (Blood) = polygenic risk scores for blood counts which were measured in the PMBB; PRS (Blood ext.) polygenic risk scores for an extended set of blood traits (see Methods for details).

**Figure S6:**
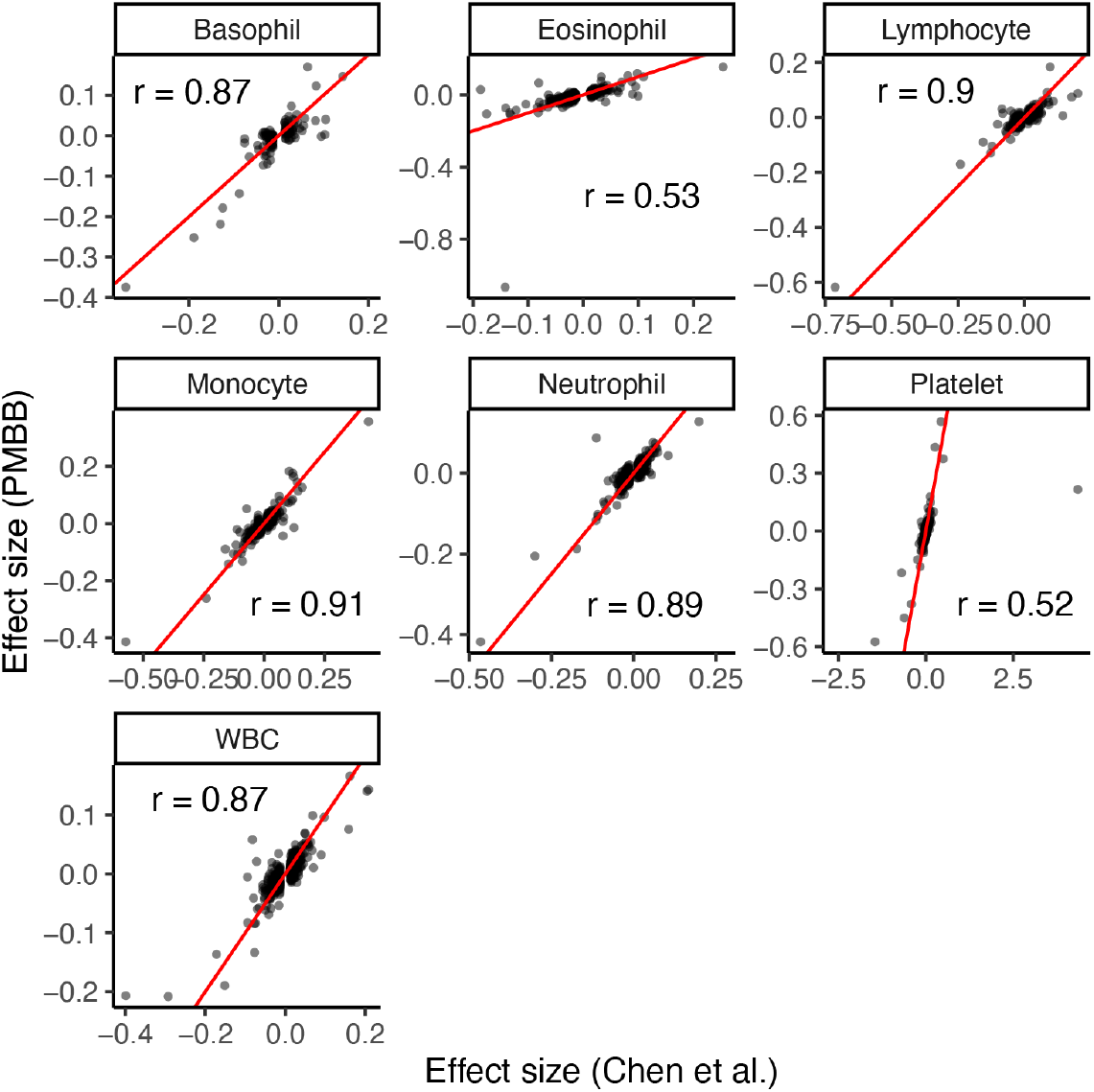
Effect sizes for variants discovered in Chen *et al*. [25] are correlated with their effects estimated in the PMBB EUR cohort. The red line represents *y* = *x*. The effects could only be re-estimated for traits which were available in the PMBB. One variant which can be seen as having a large effect size on platelet counts as estimated by Chen *et al*. was removed (see for more details). The numbers in each plot show the correlation coefficients.

**Figure S7:**
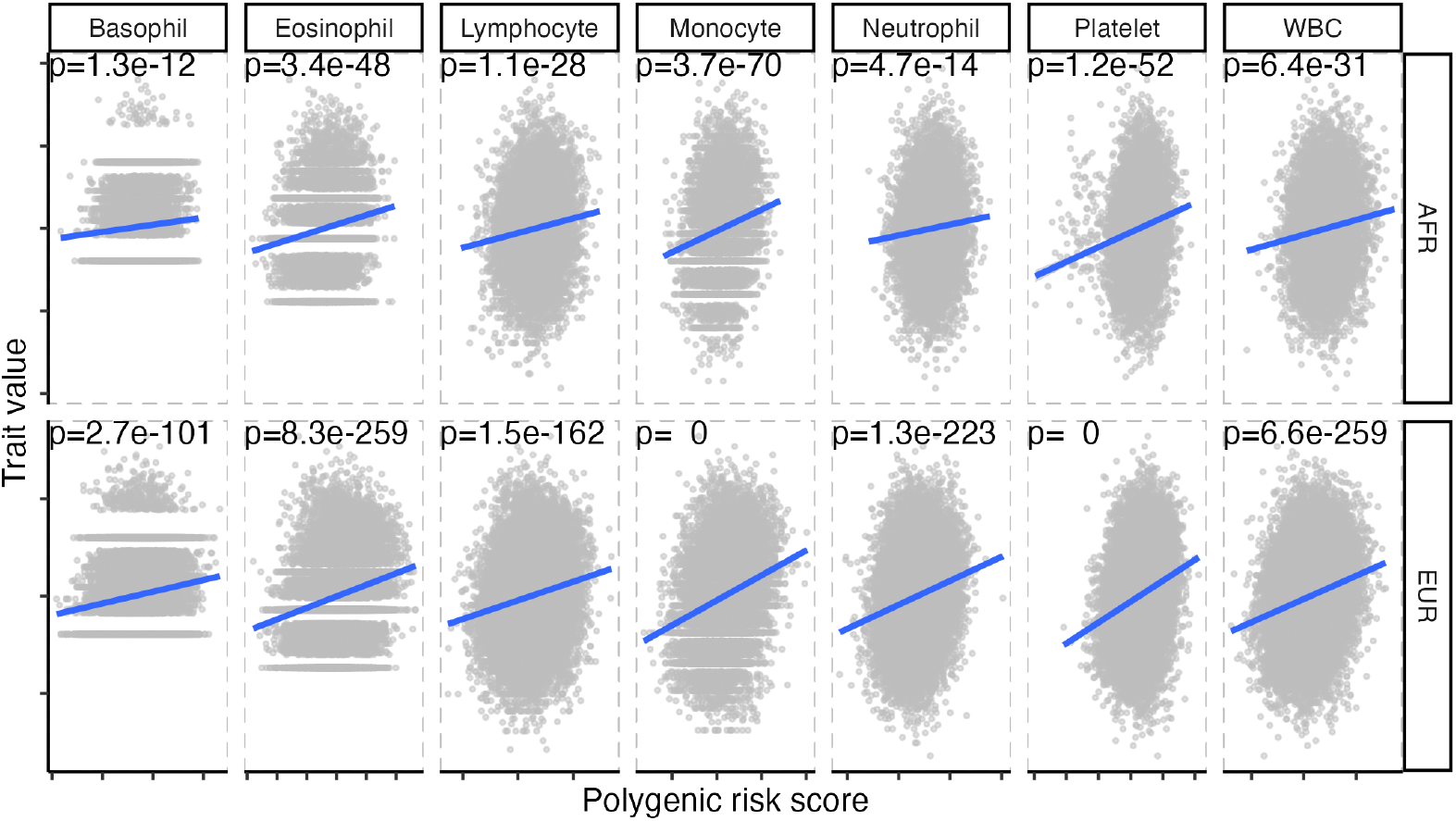
Polygenic risk scores (PRS) constructed using effects from Chen *et al*. [25] are strongly correlated with the actual phenotypes in both PMBB cohorts. The blue line represents the linear regression line.

**Figure S8:**
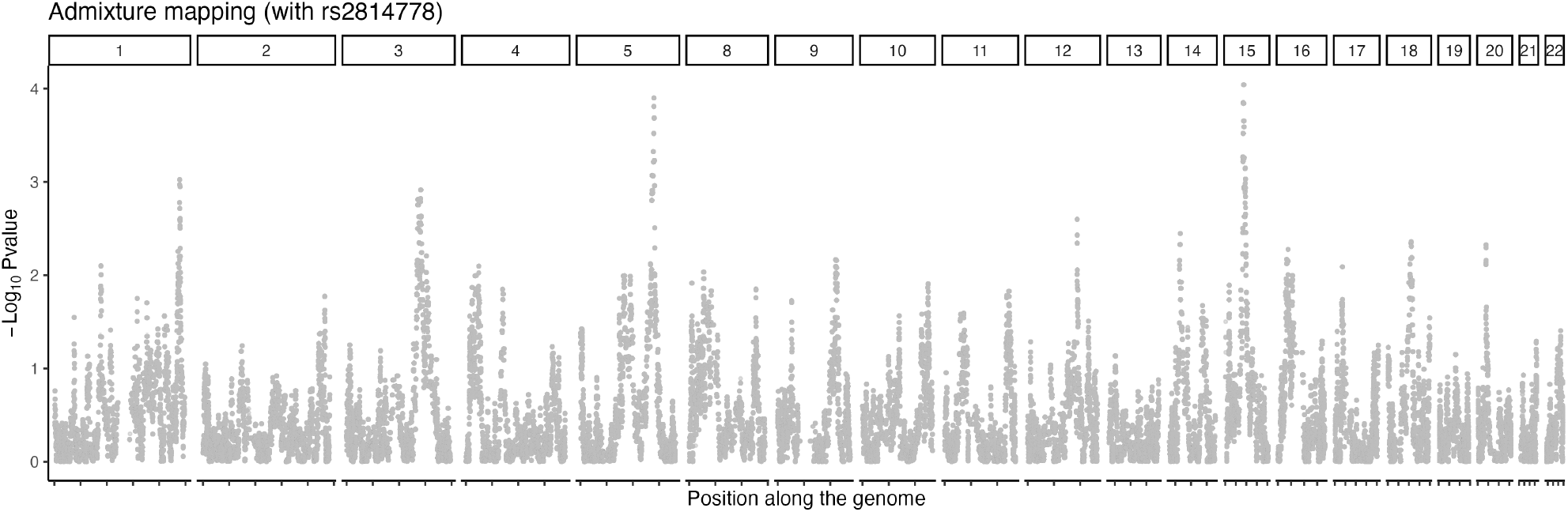
Admixture mapping of rlrmtCN in the AFR cohort. The x-axis shows the position along the genome and the y-axis shows the -log_10_ of the p-value of association between local ancestry at each position and rlrmtCN. Global ancestry proportion and the Duffy-null genotype were included as covariates.

**Figure S9:**
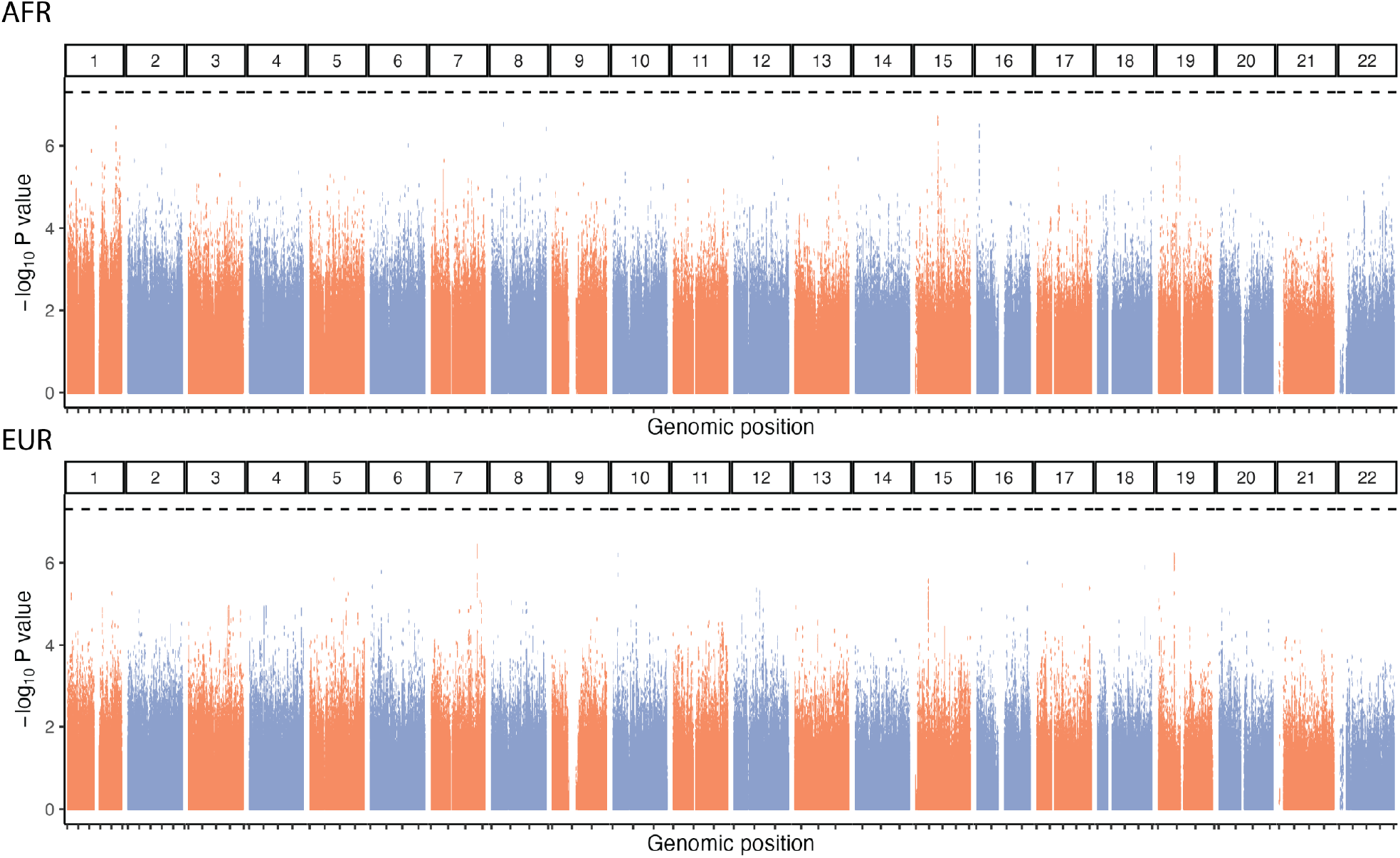
GWAS of mtDNA copy number (rlrmtCN) carried out separately in the AFR and EUR cohorts. The x-axis shows the genomic position, grouped by chromosomes (vertical panels) and the y-axis shows the -log_10_ of the association p-value. The dotted horizontal line represents the genome-wide significance threshold of 5 *×* 10^−08^. The first 20 PCs, computed separately within each cohort, were included as covariates.

**Figure S10:**
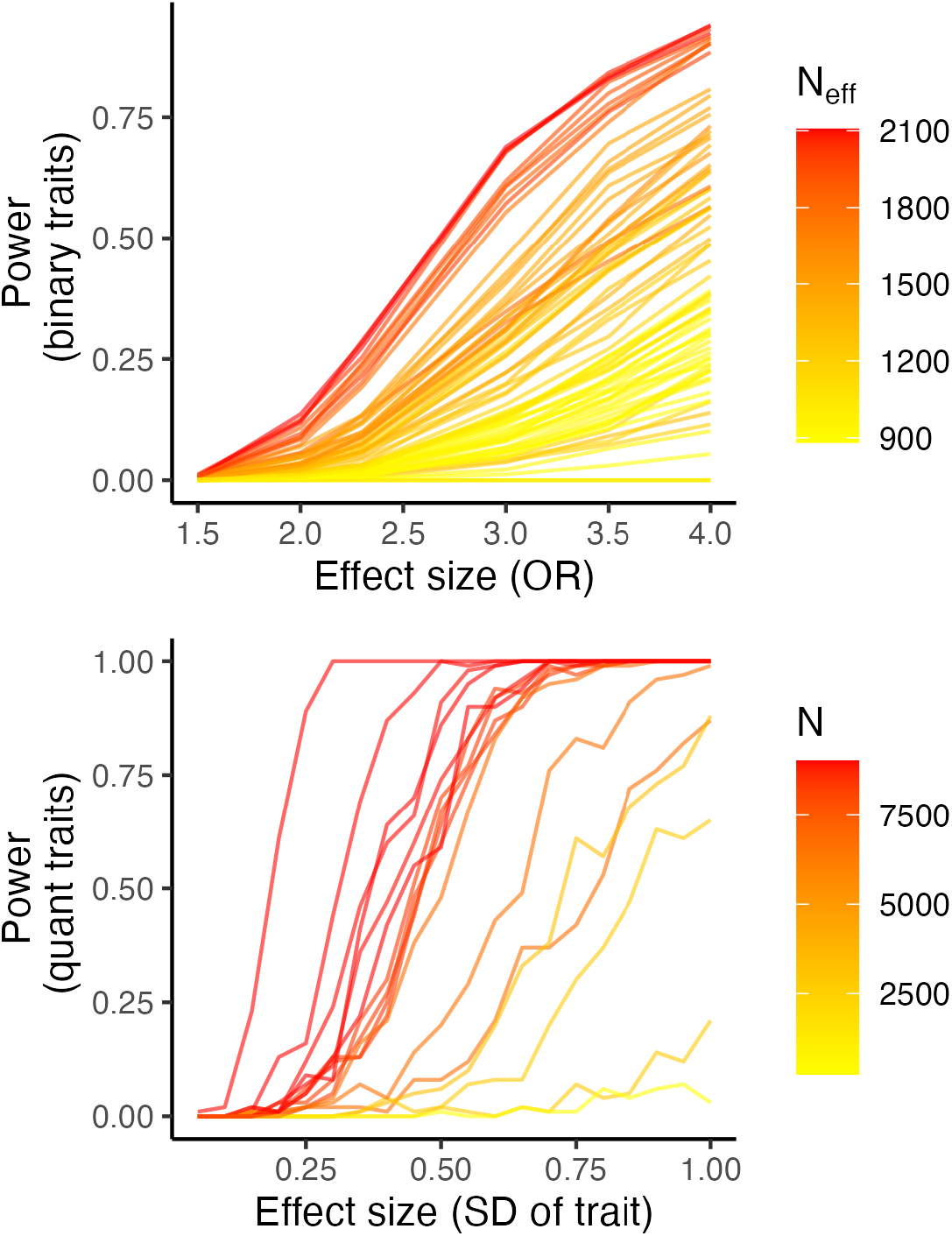
Power to detect a significant interaction effect between mitochondrial and nuclear ancestry for binary traits (case/control data) and quantitative traits (e.g. lab measurements). The x-axis lists the effect size, i.e., odds ratio (OR) or in units of standard deviation, for binary and quantitative traits, respectively and the y-axis shows the power of detecting an interaction effect at *α* = 5 *×* 10^−05^. For quantitative traits, the color represents the sample size and for binary traits, it represents the effective sample size (N_eff_): *nϕ*(1 − *ϕ*) where *ϕ* is the proportion of cases and n is the sample size.

**Figure S11:**
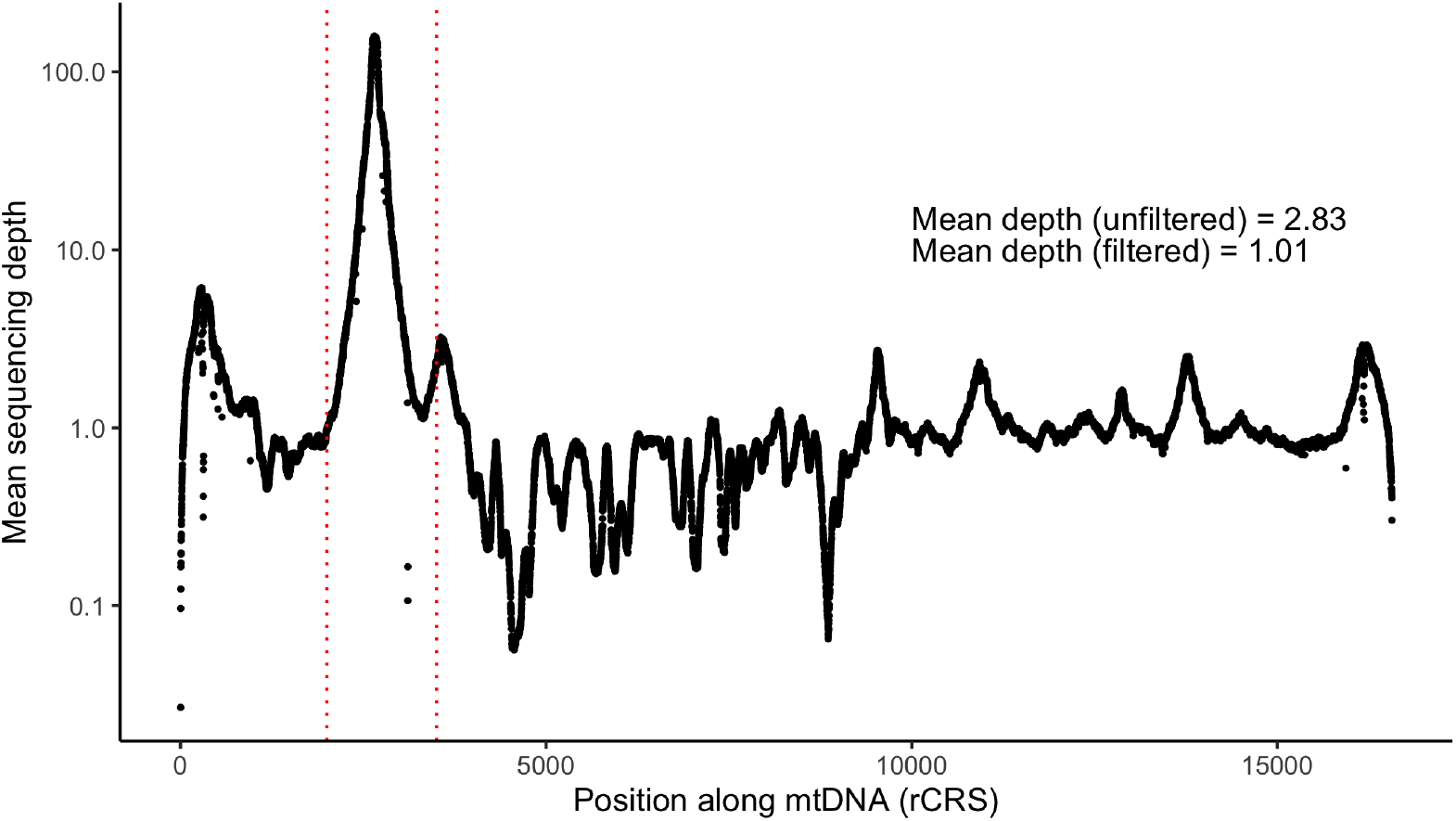
Mean sequencing depth (across individuals in the PMBB) of off-target reads aligning to the Revised Cambridge Reference Sequence (rCRS) of human mtDNA. Note that the y-axis is on a log-scale. Depth values from the region between the dotted red lines were filtered out for subsequent analysis.

## References

1. Rath, S., Sharma, R., et al. MitoCarta3.0: an updated mitochondrial proteome now with sub-organelle localization and pathway annotations. Nucleic Acids Research 49, D1541–D1547 (D1 2021).

2. Taylor, R. W. & Turnbull, D. M. Mitochondrial DNA mutations in human disease. Nature Reviews Genetics 6, 389–402 (5 2005).

3. Chinnery, P. F. Primary Mitochondrial Disorders Overview. GeneReviews® (2021).

4. D’Erchia, A. M., Atlante, A., et al. Tissue-specific mtDNA abundance from exome data and its correlation with mitochondrial transcription, mass and respiratory activity. Mitochondrion 20, 13–21 (2015).

5. Filograna, R., Mennuni, M., et al. Mitochondrial DNA copy number in human disease: the more the better? FEBS Letters 595, 976–1002 (8 2021).

6. Hägg, S., Jylhävä, J., et al. Deciphering the genetic and epidemiological landscape of mitochondrial DNA abundance. Human Genetics 140, 849–861 (2021).

7. Knez, J., Winckelmans, E., et al. Correlates of Peripheral Blood Mitochondrial DNA Content in a General Population. American Journal of Epidemiology 183, kwv175 (2015).

8. Picard, M. Blood mitochondrial DNA copy number: What are we counting? Mitochondrion 60, 1–11 (2021).

9. Howes, R. E., Patil, A. P., et al. The global distribution of the Duffy blood group. Nature Communications 2011 2:1 2, 1–10 (1 2011).

10. Reich, D., Nalls, M. A., et al. Reduced Neutrophil Count in People of African Descent Is Due To a Regulatory Variant in the Duffy Antigen Receptor for Chemokines Gene. PLoS Genetics 5 (ed Visscher, P. M.) e1000360 (2009).

11. Ng, Y. S. & Turnbull, D. M. Mitochondrial disease: genetics and management. Journal of Neurology 263, 179–191 (1 2016).

12. Parikh, S., Goldstein, A., et al. Diagnosis and management of mitochondrial disease: A consensus statement from the Mitochondrial Medicine Society. Genetics in Medicine 17, 689–701 (9 2015).

13. Suomalainen, A. & Battersby, B. J. Mitochondrial diseases: The contribution of organelle stress responses to pathology. Nature Reviews Molecular Cell Biology 19, 77–92 (2 2018).

14. Russell, O. M., Gorman, G. S., et al. Mitochondrial Diseases: Hope for the Future. Cell 181, 168–188 (1 2020).

15. Agarwal, S., Fulgoni, V. L., et al. Assessing alcohol intake & its dosedependent effects on liver enzymes by 24-h recall and questionnaire using NHANES 2001-2010 data. Nutrition Journal 15, 1–12 (1 2016).

16. Mansouri, A., Gaou, I., et al. An alcoholic binge causes massive degradation of hepatic mitochondrial DNA in mice. Gastroenterology 117, 181–190 (1 1999).

17. Zhang, Y., Guallar, E., et al. Association between mitochondrial DNA copy number and sudden cardiac death: Findings from the Atherosclerosis Risk in Communities study (ARIC). European Heart Journal 38, 3443–3448 (2017).

18. Wiersma, M., van Marion, D. M., et al. Cell-Free Circulating Mitochondrial DNA: A Potential Blood-Based Marker for Atrial Fibrillation. Cells 9 (5 2020).

19. Ashar, F. N., Zhang, Y., et al. Association of mitochondrial DNA copy number with cardiovascular disease. JAMA Cardiology 2, 1247–1255 (2017).

20. El-Hattab, A. W. & Scaglia, F. Mitochondrial DNA Depletion Syndromes: Review and Updates of Genetic Basis, Manifestations, and Therapeutic Options. Neurotherapeutics 10, 186–198 (2 2013).

21. Yang, J., Lee, S. H., et al. GCTA: a tool for genome-wide complex trait analysis. American journal of human genetics 88, 76–82 (1 2011).

22. Longchamps, R. J., Yang, S. Y., et al. Genome-wide analysis of mitochondrial DNA copy number reveals loci implicated in nucleotide metabolism, platelet activation, and megakaryocyte proliferation. Human Genetics 141, 127–146 (2022).

23. Genovese, G., Friedman, D. J., et al. Association of trypanolytic ApoL1 variants with kidney disease in African Americans. Science 329, 841–5 (2010).

24. Limou, S., Nelson, G. W., et al. APOL1 kidney risk alleles: population genetics and disease associations. Adv Chronic Kidney Dis 21, 426–33 (2014).

25. Chen, M. H., Raffield, L. M., et al. Trans-ethnic and Ancestry-Specific Blood-Cell Genetics in 746,667 Individuals from 5 Global Populations. Cell 182, 1198–1213.e14 (5 2020).

26. Cai, N., Li, Y., et al. Genetic Control over mtDNA and Its Relationship to Major Depressive Disorder. Current Biology 25, 3170–3177 (24 2015).

27. Chong, M., Mohammadi-Shemirani, P., et al. GWAS and ExWAS of blood Mitochondrial DNA copy number identifies 71 loci and highlights a potential causal role in dementia. eLife 11 (2022).

28. Zaidi, A. A. & Makova, K. D. Investigating mitonuclear interactions in human admixed populations. Nature Ecology and Evolution 3, 213–222 (2019).

29. Crawford, N., Prendergast, D., et al. Divergent Patterns of Mitochondrial and Nuclear Ancestry Are Associated with the Risk for Preterm Birth. The Journal of Pediatrics 194, 40–46.e4 (2018).

30. Udler, M. S., Nadkarni, G. N., et al. Effect of genetic African ancestry on EGFR and kidney disease. Journal of the American Society of Nephrology 26, 1682–1692 (2015).

31. Zilbermint, M., Hannah-Shmouni, F., et al. Genetics of Hypertension in African Americans and Others of African Descent. International Journal of Molecular Sciences 20, 1081 (2019).

32. Samokhvalov, A. V., Irving, H., et al. Alcohol consumption, unprovoked seizures, and epilepsy: A systematic review and meta-analysis. Epilepsia 51, 1177–1184 (7 2010).

33. Wu, L. T., Woody, G. E., et al. Racial/Ethnic Variations in Substance-Related Disorders Among Adolescents in the United States. Archives of General Psychiatry 68, 1176–1185 (11 2011).

34. Holmbeck, M. A., Donner, J. R., et al. A Drosophila model for mito-nuclear diseases generated by an incompatible interaction between tRNA and tRNA synthetase. DMM Disease Models and Mechanisms 8, 843–854 (8 2015).

35. Burton, R. S., Ellison, C. K., et al. The sorry state of F2 hybrids: Consequences of rapid mitochondrial DNA evolution in allopatric populations in. 168 (The University of Chicago Press, 2006).

36. Meiklejohn, C. D., Holmbeck, M. A., et al. An Incompatibility between a Mitochondrial tRNA and Its Nuclear-Encoded tRNA Synthetase Compromises Development and Fitness in Drosophila. PLoS Genetics 9 (ed Payseur, B. A.) e1003238 (1 2013).

37. Reinhardt, K., Dowling, D. K., et al. Medicine. Mitochondrial replacement, evolution, and the clinic. Science 341, 1345–6 (2013).

38. Eyre-Walker, A. Mitochondrial Replacement Therapy: Are Mito-nuclear Interactions Likely To Be a Problem? Genetics 205, 1365–1372 (4 2017).

39. Rausser, S., Trumpff, C., et al. Mitochondrial phenotypes in purified human immune cell subtypes and cell mixtures. eLife 10 (2021).

40. Denny, J. C., Ritchie, M. D., et al. PheWAS: demonstrating the feasibility of a phenome-wide scan to discover gene–disease associations. Bioinformatics 26, 1205–1210 (9 2010).

41. Bastarache, L. Using Phecodes for Research with the Electronic Health Record: From PheWAS to PheRS. https://doi.org/10.1146/annurev-biodatasci-122320-112352 4, p1–19 (1 2021).

42. García-Olivares, V., Muñoz-Barrera, A., et al. A benchmarking of human mitochondrial DNA haplogroup classifiers from whole-genome and whole-exome sequence data. Scientific Reports 11, 1–11 (2021).

43. Fast two-stage phasing of large-scale sequence data. American Journal of Human Genetics 108, 1880–1890 (2021).

44. Maples, B. K., Gravel, S., et al. RFMix: a discriminative modeling approach for rapid and robust local-ancestry inference. American journal of human genetics 93, 278–88 (2013).

45. Auton, A., Abecasis, G. R., et al. A global reference for human genetic variation. Nature 2015 526:7571 526, 68–74 (7571 2015).

46. Alexander, D. H., Novembre, J., et al. Fast model-based estimation of ancestry in unrelated individuals. Genome research 19, 1655–64 (2009).

47. Das, S., Forer, L., et al. Next-generation genotype imputation service and methods. Nature Genetics 48, 1284–1287 (10 2016).

48. Chang, C. C., Chow, C. C., et al. Second-generation PLINK: rising to the challenge of larger and richer datasets. GigaScience 4, s13742.#x2013;015–0047–8 (1 2015).

49. Shriner, D., Adeyemo, A., et al. Joint Ancestry and Association Testing in Admixed Individuals. PLOS Computational Biology 7, e1002325 (12 2011).

50. Plummer, M., Best, N., et al. CODA: Convergence Diagnosis and Output Analysis for MCMC. R News 6, 7–11 (2006).

51. R Core Team. R: A Language and Environment for Statistical Computing R Foundation for Statistical Computing (Vienna, Austria, 2021).

